# Tricycle surveillance in Antananarivo, Madagascar: circulation of both extended-spectrum beta-lactamase producing *Escherichia coli* strains and plasmids among humans, chickens and the environment

**DOI:** 10.1101/2023.01.16.23284583

**Authors:** Milen Milenkov, Caroline Proux, Tiavina Lalaina Rasolofoarison, Fetra Angelot Rakotomalala, Saida Rasoanandrasana, Lalaina Vonintsoa Rahajamanana, Christian Rafalimanana, Zakasoa Ravaoarisaina, Ilo Tsimok’Haja Ramahatafandry, Emilie Westeel, Marie Petitjean, Julie Marin, Jimmy Mullaert, Lien Han, Olivier Clermont, Laurent Raskine, Hubert Endtz, Antoine Andremont, Erick Denamur, Florence Komurian-Pradel, Luc Hervé Samison, Laurence Armand-Lefevre

## Abstract

**Background:** Antimicrobial resistance is a major public health threat, affecting not only humans but also animals and the environment. Although the “One Health” dimension of resistance is well recognized, data are lacking on the circulation of resistance, particularly in low-income countries. The World Health Organization has proposed a protocol called Tricycle, focusing on extended-spectrum beta-lactamase (ESBL)-*Escherichia coli* surveillance in the three sectors. We implemented Tricycle in Madagascar to assess ESBL-*E. coli* prevalence and describe intra- and inter-sector circulation of ESBL-*E. coli* and plasmids.

**Methods:** 289 pregnant women, 246 farm chickens and 28 surface waters were sampled in Antananarivo (the capital city) area and tested for ESBL-*E. coli*. Isolates were sequenced by short-(Illumina) and long-(Nanopore) read methods.

**Findings:** ESBL-*E. coli* prevalence was 29·8%, 56·9% and 100% in pregnant women, chickens, and the environment, respectively. The wet season was associated with higher rates of carriage in humans (OR=3·1, 95%CI 1·8-5·3) and animals (OR=2·8 95%CI 1·7-4·8). Sequencing of 277 non-duplicated isolates (82, 118 and 77 from each sector, respectively) showed high genetic diversity (90 STs identified) with differences between sectors. Single nucleotide polymorphism (SNP) analysis revealed 169/277 (61%) isolates grouped into 44 clusters (≥2 isolates) of closely related isolates (<40 SNPs), of which 24 contained isolates from two sectors and five contained isolates from all three sectors. ESBL genes were all *bla*_CTX-M_ (77.6% *bla*_CTX-M-15_), chromosomally integrated in 57·4% (159/277) of isolates, and plasmidic in 40·8% (113/277). The 114 ESBL-carrying plasmids were mainly IncF (55·2%, n=63) and IncY (36·8%, n=42). The F31/36:A4:B1 (n=13) and F-:A-:B53 (n=8) subtypes, and all IncY plasmids, highly conserved, were observed in isolates of differing genetic backgrounds from all sectors.

**Interpretation:** Despite varying strain population structures in the three sectors, both ESBL-*E. coli* strains and plasmids are circulating among humans, chickens and the environment in the capital of Madagascar.

**Funding:** Fondation Mérieux, INSERM, Université Paris Cité

## Introduction

For decades, antimicrobial resistance (AMR) was considered exclusively from a human health point of view, but has now come to be regarded as a global problem also involving animal health, food production and environmental conditions.^1^ Scientists now agree on the importance of a holistic One Health approach with epidemiological surveillance as a key element in the fight against AMR.^2^ In this context, the World Health Organization (WHO) has developed a simplified, integrated, multisectoral surveillance initiative called Tricycle, using a One Health approach in order to address resistance in three major sectors: human (community carriage and bloodstream infections), the food-chain (live poultry) and the environment (surface water, slaughterhouse effluents and wastewater).^3^ The Tricycle protocol focuses on a single key indicator: the prevalence of Extended-Spectrum Beta-Lactamase (ESBL) producing *Escherichia coli*. ESBL-*E. coli* was chosen as the target organism because ESBL producing *Enterobacterales* are viewed as one of the biggest threats in terms of AMR, particularly as it is one of the species most often isolated from invasive infections in humans.^4^ In addition, *E. coli* is a commensal of the human gut, a wide variety of warm-blooded animals, and is also widespread in the environment.^5,6^ AMR surveillance studies, aimed at investigating links between resistant bacteria in humans, animals and the environment on a same temporal and geographical scale, are still rare. Moreover, such studies rarely include the investigation of resistance circulation via plasmids.

The Tricycle protocol was implemented in 2018 in Antananarivo, Madagascar. Here, we report the results of the first year of the Tricycle project describing the prevalence of ESBL-*E. coli* in healthy pregnant women, in the food-chain and in the environment. We thus took advantage of a strong epidemiological collection to compare the genomic characteristics of the strains isolated from the three sectors, and to investigate the intra- and inter-sectoral circulation of ESBL-*E. coli* and ESBL-carrying plasmids.

## Methods

### Study design and setting

Sampling and microbiological methods were performed according to the Tricycle protocol.^3^ Human isolates are part of a larger collection of ESBL-*E. coli* strains from Malagasy pregnant women, described previously.^7^ In brief, healthy pregnant women were enrolled from July 2018 to April 2019, after having provided written informed consent. Rectal swabs were collected in three maternity wards in Antananarivo (Joseph Raseta Befelatanana hospital [JRB], Mère-Enfant Tsaralalana hospital [TSA], and Joseph Ravoahangy Andrianavalona hospital [JRA]) and sent to each hospital laboratory. Animal and environmental samples were collected from May 2018 to April 2019 and processed at the Charles Mérieux Center of Infectious Disease (CICM) in Antananarivo. Live chickens were purchased in batches of six per week from 27 different wet markets (41 batches) in Antananarivo, with chickens from the same batch coming from the same farm and kept in the same cage during transport. Environmental sampling was conducted every two months (7 sampling campaigns) on surface water from Ikopa river, which crosses the city. Samples were collected at sites upstream (5·5 km from downtown), downstream (5·4 km from downtown), from a wastewater channel (in the city center), and from a slaughterhouse sewage (5·4 km from downtown) (**Figure S1**).

### Microbiology

Rectal swabs and chicken caeca were plated on MacConkey agar with 4 mg/L cefotaxime. Environmental samples were plated quantitatively on TBX agar without and with 4 mg/L cefotaxime. For each positive plate, three (human and animal) or five (environment) ESBL-*E. coli* presumptive colonies were selected and tested for indole production. ESBL production was confirmed by double disc synergy test on antibiotic susceptibility testing. ^3^ For environmental sampling, total *E. coli* and ESBL-*E. coli* colonies were counted and respective concentrations calculated. Strains were shipped to France where an identification and an extended antibiogram (32 antibiotics) were performed.

### Whole genome sequencing

#### Illumina technology

For each sample, all isolates with distinct antimicrobial susceptibility profiles, were selected for whole genome sequencing (WGS) (see appendix).

#### Nanopore technology

Isolates carrying at least one plasmid replicon were also sequenced using Nanopore technology. DNA extraction was performed using NucleoMag 96 Tissue kit (Macherey Nagel, Hœrdt, France) and sequencing was performed on a MinION platform (Oxford Nanopore Technologies, Oxford, UK).

### Bioinformatics

Plasmid sequences were reconstructed using Illumina and Nanopore reads with Unicycler v0.4.9b software.^8^ Further details of laboratory methods including genome assembly, antibiotic resistance and virulence gene detection, replicon detection, phylogenetic group, serotype, multilocus sequence typing (MLST), phylogenetic analysis, as well as isolate genome and plasmid comparison are available in the appendix.

### Statistical analysis

After sequencing, to avoid over-representation of clonal strains in chicken samples, similar isolates (<10 SNPs) from different chickens within the same batch were considered as duplicates and removed from the final analysis.

Factors associated with ESBL-*E. coli* carriage were identified with univariable logistic regressions with carriage as dependent variable. Proportion of each gene from different sectors was compared by Fisher exact test, and the reported p-value was corrected for multiple comparisons by the Benjamini-Hochberg method. Further details regarding statistical analysis are available in the appendix.

### Ethics

The study was approved by the Ethics Committee of Ministry of Health, Madagascar (038-MSANP/CERBM).

## Results

### *Prevalence and concentration of ESBL-*E. coli *and associated risk factors*

In all, 289 pregnant women, 246 chickens and 28 environmental samples were included in the study (**Table S1**). ESBL-*E. coli* prevalence was 29·8% (86/289) in pregnant women and 56·9% (140/246) in chickens. All environmental samples contained ESBL-*E. coli* (100%, 28/28), with concentration ranging from 1·11 logCFU/100ml in upstream surface water, to 2·37 in downstream water, 4·06 in wastewater and 4·09 in slaughterhouse effluents (p<0·0001)(**Figure S2**).

The only factor associated with ESBL-*E. coli* carriage was a sampling during the wet and warm season in both humans (OR=1·5 per 100mm rainfall and OR=1·3 per °C, p<0·001) and chickens (OR=1· 4 per 100mm rainfall and OR=1·2 per °C, p<0·0001) (**Table 1**). Variations in ESBL-*E. coli* concentrations in the environment were not related to seasonality.

**Table 1.** Risk factors associated with ESBL-*E. coli* carriage in pregnant women and chickens

| Sector | Variable |  | N non carriers<br>(%)<br>or Mean (SD) | N carriers<br>(%)<br>or Mean (SD) | OR (95%CI) | p |
| --- | --- | --- | --- | --- | --- | --- |
| Human | N = 289 |  | 203 | 86 |  |  |
|  |  | <20 | 22 (10.9) | 9 (10.5) | 1 (ref) |  |
|  | Age by category<br>(missing = 1) | 21-25 | 67 (33.2) | 31 (36.0) | 1.13 (0.48 - 2.85) | 0.78505 |
|  |  | 26-30 | 72 (35.6) | 26 (30.2) | 0.88 (0.37 - 2.24) | 0.7849 |
|  |  | >30 | 41 (20.3) | 20 (23.3) | 1.19 (0.47 - 3.16) | 0.71423 |
|  | Age Mean (SD)<br>(missing=1) |  | 26.57 (5.57) | 26.88 (5.87) | 1.01 (0.97 - 1.06) | 0.66517 |
|  | Season | Wet vs. Dry | 48 (23.6) | 42 (48.8) | 3.08 (1.81 - 5.27) | **0.00003** |
|  | Rainfall Mean (SD)<br>OR per 100mm |  | 47.78 (93.48) | 98.15 (121.82) | 1.53 (1.21 - 1.96) | **0.00044** |
|  | Temperature (°C)<br>Mean (SD) |  | 18.67 (1.96) | 19.59 (1.95) | 1.26 (1.11 - 1.44) | **0.00042** |
|  | Education | Absent or elementary | 12 (5.9) | 6 (7.0) | 1 (ref) |  |
|  |  | Secondary | 108 (53.2) | 41 (47.7) | 0.76 (0.28 - 2.3) | 0.60507 |
|  |  | University or graduate | 83 (40.9) | 39 (45.3) | 0.94 (0.34 - 2.87) | 0.90778 |
|  | Private Toilets |  | 101 (49.8) | 45 (52.3) | 1.11 (0.67 - 1.84) | 0.68934 |
|  | Electricity access |  | 189 (93.1) | 80 (93.0) | 0.99 (0.38 - 2.87) | 0.98041 |
|  | Drinking water access | Private | 61 (30.0) | 19 (22.1) | 1 (ref) |  |
|  |  | Shared | 142 (69.9) | 67 (77.9) | 1.32 (0.72 - 2.5) | 0.38229 |
|  | Pets or farm animals |  | 95 (46.8) | 34 (39.5) | 0.74 (0.44 - 1.24) | 0.25681 |
|  | Recent antibiotics intake |  | 23 (11.3) | 13 (15.1) | 1.39 (0.65 - 2.86) | 0.37439 |
| Chicken | N = 246 |  | 106 | 140 |  |  |
|  | Weight (g) mean (SD) |  | 804.11 (155.86) | 788.57 (164.57) | 1 (1 - 1) | 0.45267 |
|  | Season | Wet vs. dry | 31 (29.0) | 74 (53.2) | 2.79 (1.65 - 4.81) | **0.00017** |
|  | Rainfall mean (SD)<br>OR per 100mm |  | 66.41 (142.63) | 179.74 (216.77) | 1.4 (1.21 - 1.65) | **0.00002** |
|  | Temperature (°C)<br>Mean (SD) |  | 18.77 (2.71) | 20.24 (2.63) | 1.22 (1.11 - 1.35) | **0.00005** |

### Strain characteristics and comparison by sector

Among the 672 isolates received in France, 654 were subjected to identification and antibiotic susceptibility testing. Then, 296 isolates with non-similar resistance patterns were selected for WGS, among which 19, considered as duplicates, were excluded. In all, 277 isolates (82 from human, 118 from animal and 77 from the environment) from 213 samples were included for further analysis (**Figure S3**).

#### Global comparison

Analysis of the 277 *E. coli* genomes revealed a core-genome of 2,522,760 bp containing 2623 genes. Pangenome calculations indicated that *E. coli* genomic diversity represents an open pangenome model containing a reservoir of more than 17,200 genes. Comparison of the core and pan genomes showed quicker decay in the pangenomic gene content of chicken isolates than of human and environmental ones. The genetic diversity of chicken isolates was lower than that of human isolates, which was in turn lower than that of environmental isolates (p<2^-16^). The mean genome size of chicken isolates (4·77 Mbp) was smaller than that of human isolates (4·82 Mbp) and of environmental isolates (4·92 Mbp) (p<10^−7^). Inter-sectoral genetic diversity was higher than intra-sectoral genetic diversity (p<2·2^-16^) (**Figure S4**).

#### Phylogroup and sequence types

Isolates from commensalism-associated phylogenetic groups A and B1 were predominant in all sectors, accounting for 91·5% (108/118) of chicken, 86·6% (71/82) of human and 70·1% (54/77) of environmental isolates (p<0·01) (**Figure S5**). Isolates were highly diverse as MLST identified 90 different STs among the 277 isolates including 48 STs in 118 chicken, 52 in 82 human, and 39 in 77 environmental isolates (p<0·05). Among them, 22, 20, and 10 STs were observed, respectively, in animal, human and environmental sectors only, whereas 27 STs were identified in two sectors and 11 STs in all three sectors (**Figure 1)**. The extra-intestinal, virulent-associated, B2 phylogenetic group-ST131 was detected only in human and environmental sectors. The clonal complex STc10 was the most prevalent in the three sectors, impacting 45·8% (54/118), 35·4% (29/82), and 31· 2% (24/77) of chicken, human, and environmental isolates, respectively (p=0·29). Interestingly, the four most frequent STc identified in the human sector, STc10, STc46, STc155 and ST450 (no STc), accounting for 60% of human isolates, were detected in all three sectors (**Figure S6**). A phylogenetic tree, based on the SNPs core-genome illustrates the high genetic diversity of ESBL *E. coli* isolates (**Figure 2)**.

**Figure 1.**
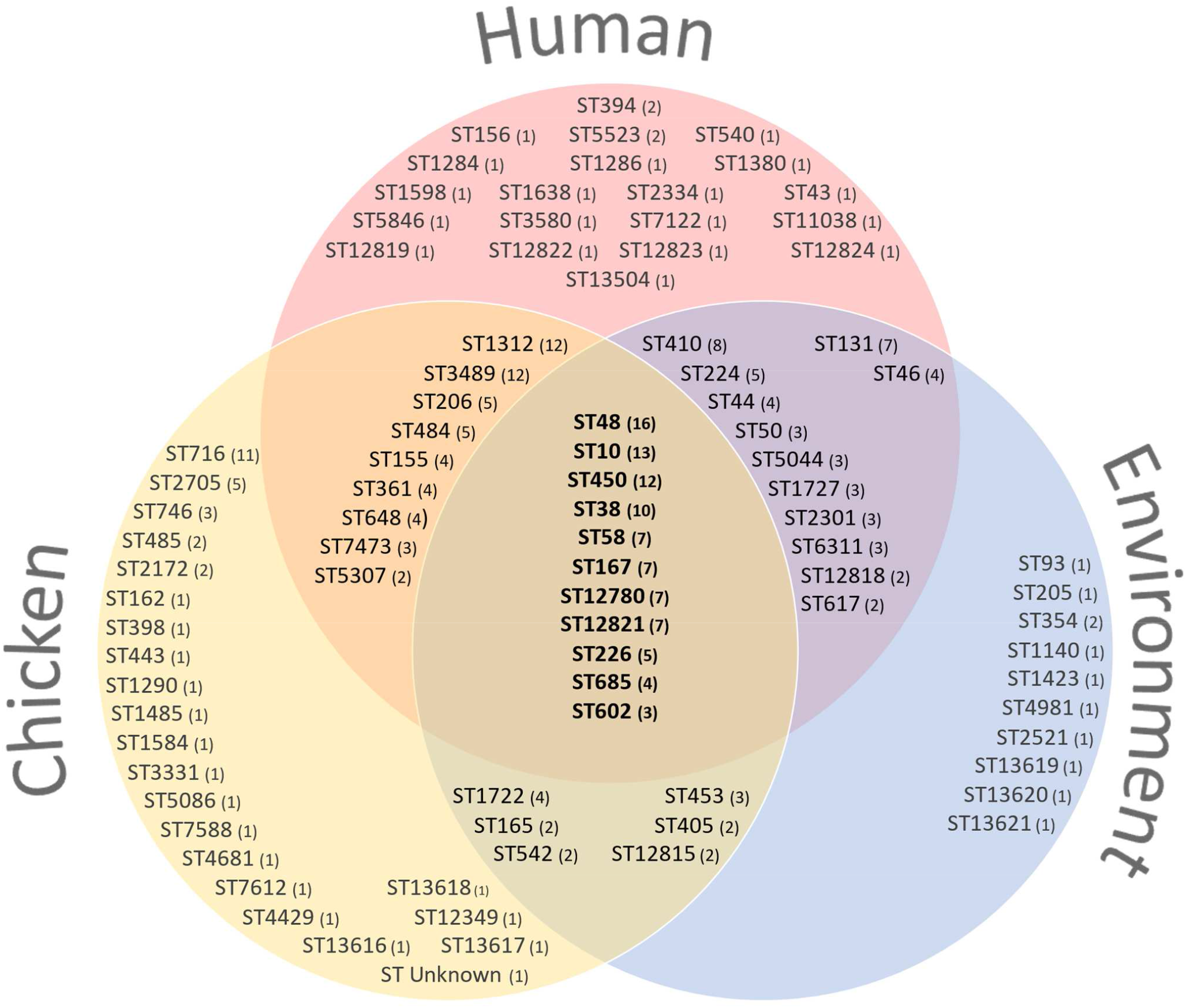
Venn diagram of sequence types (ST) distribution between the three sectors. Numbers in brackets indicate the number of isolates belonging to the respective ST.

**Figure 2.**
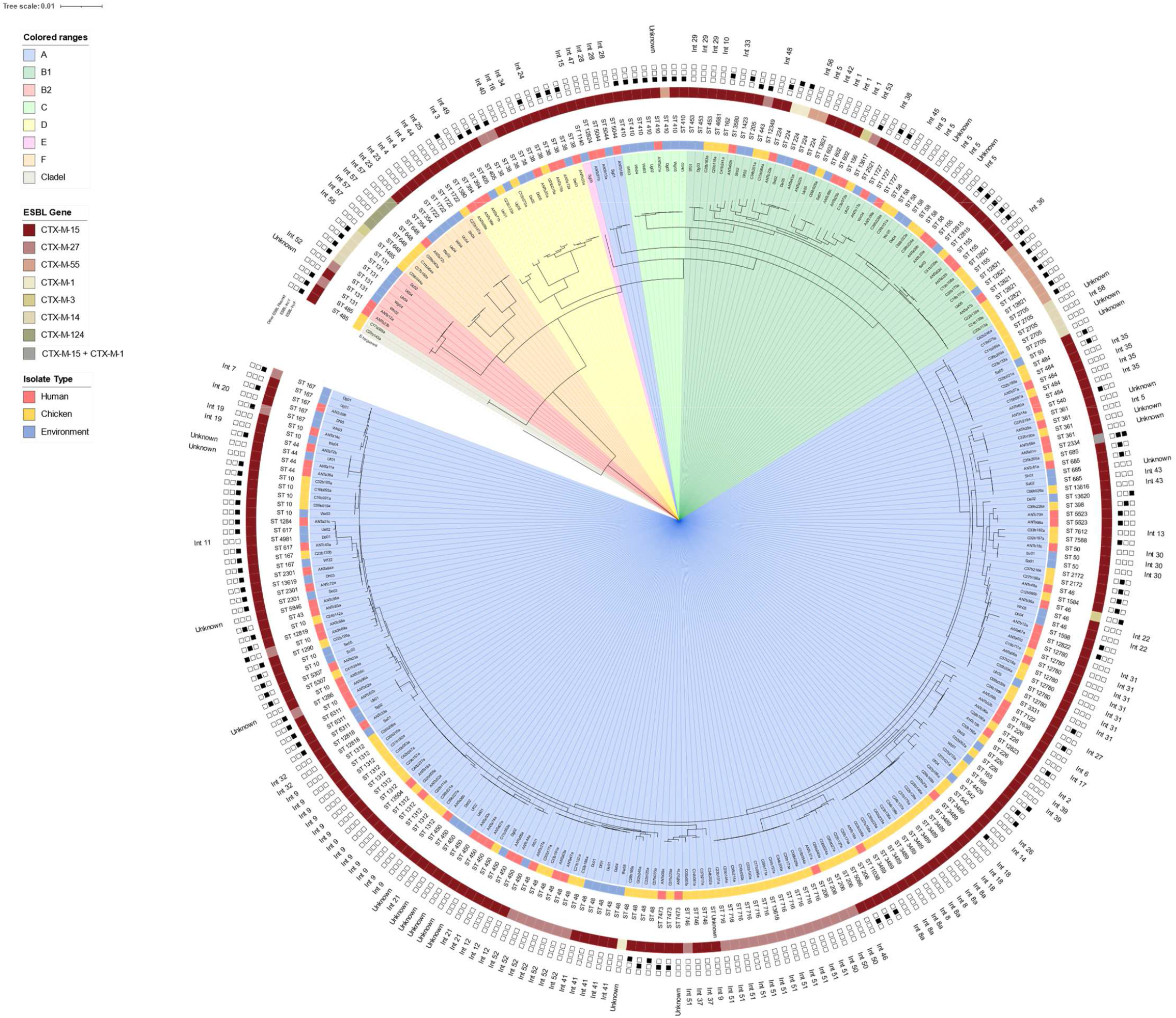
Phylogenetic tree based on core genome sequences of the 277 ESBL-*E. coli* isolates. Isolates names are highlighted depending on their phylogenetic group (inner ring). Colored strips represent isolates origin (ring two) and the CTX-M enzyme detected (ring four). STs are given in the third ring. The two last rings report the localization of the *bla*_CTX-M_ genes: the penultimate ring represents the type of ESBL carrying plasmids and the outer ring represents the chromosomal integration site of the *bla*_CTX-M_ gene, determined by the upstream and downstream CDS surrounding the of *bla*_CTX-M_ gene. Correspondences between integration site code and respective upstream and downstream CDS are listed in Table S2. “Unknown” corresponds to isolates with a chromosomally integrated *bla*_CTX-M_ gene, but with no integration site identified, because of *bla*_CTX-M_ contigs too short to identify the *bla*_CTX-M_ genetic environment.

#### Antibiotic resistance and resistance genes

All isolates were highly resistant to all beta-lactams, except to piperacillin/tazobactam (13·7%), cefoxitin (6·1%) and imipenem (0·7%). They were frequently resistant to fluoroquinolones (ciprofloxacin [76·9%]), co-trimoxazole (63·2%) and less to aminoglycosides (gentamicin [39·7%] and amikacin [9·7%]) with slight variation between sectors (**Figure S7A**). One chicken and one environmental isolates were resistant to imipenem.

The mean number of resistance genes per isolate was higher in the environment (n=8·2) than in the human (n=7·8) and animal sectors (n=6·2), (p<0·001) (**Figure S4B**). In all isolates, the ESBL phenotype was due to *bla*_CTX-M_ genes, the most prevalent being *bla*_CTX-M-15_ (77·6%, 215/277), followed distantly by *bla*_CTX-M-27_ (12·3%, 34/277), with variation between sectors (**Figure S8**). Prevalence of other resistance genes varied also slightly among sectors (**Figure S7B**). The carbapenem resistant chicken isolate carried the *bla*_NDM-5_ gene and the environmental one co-carried the *bla*_NDM-5_ and *bla*_OXA-181_ genes.

#### Plasmid content and ESBL location

The mean number of plasmid replicons was higher in environmental (n=2·8) than in human (n=1·8) and in chicken (n 1·3) isolates (p<10^−6^) **(Figure S4B**). IncF was the most prevalent replicon in all sectors (30·5% of chicken, 50% of human and 70·1% of environmental isolates, p<0·05), followed by IncY in humans (25·6%) and chickens (22· 9%), or IncI in the environment (23· 4%).

The *bla*_CTX-M_ gene was located on the chromosome in 57·4% (159/277) of the isolates without difference between sectors, and on a plasmid in 40·8% (113/277), including two isolates with both locations (chromosomic + plasmidic on IncF-*bla*_CTX-M-15_) and one isolate with two ESBL carrying plasmids (IncN-*bla*_CTX-M-1_ and IncY-*bla*_CTX-M-15_). The location of *bla*_CTX-M_ could not be confirmed in seven isolates. Chromosomal integration rate was 56·7% for *bla*_CTX-M-15_ and 70·6 % for *bla*_CTX-M-27_ (p=0.26) and did not differ among sectors. At least 58 different integration sites were identified, located mostly in the regions corresponding to 0-65 minutes of the *E. coli* K-12 linkage map (**Figure S9**).^9^ The 114 ESBL carrying plasmids were mostly IncF (55·3%, n=63) and IncY (36·8%, n=42). Other ESBL-plasmids were IncN (n=3), IncI and IncB/O/K/Z (n=2 each), IncP and p0111 (n=1 each). Prevalence of ESBL-IncF and ESBL-IncY plasmids was 18·6% (n=22) and 14·4% (n=17), respectively, in chicken isolates, 20·7% (n=17) and 22% (n=18) in human isolates, and 31·2% (n=24) and 9·1% (n=7) in environmental isolates with no significant difference between sectors (**Figure S7D**). ESBL-IncF plasmids harboured various *bla*_CTX-M_ genes (68·3% *bla*_CTX-M-15_, 12·7% *bla*_CTX-M-55_ and *bla*_CTX-M-27_ and 6·3% *bla*_CTX-M-14_), while ESBL-IncY harboured exclusively *bla*_CTX-M-15_ (**Figure S10**).

#### Virulence associated genes (VAG)

A total of 65 different VAG clusters/operons was detected in the 277 isolates. The mean number of VAG clusters per isolate was significantly higher in environmental (n=21·1) than in human (n=19·9) and in chicken isolates (n=18·7) (p<0.0001) **(Figures S4B**). Globally, isolates carried mostly adhesins (34· 1%), genes involved in iron acquisition (33·2%), protectins/invasins (25·4%), and to a much lesser extent toxins (6.2%), with slight variation among sectors **(Figures S7C**).

### Strain circulation

We first investigated the intra- and inter-sector strain circulation. The global phylogenetic tree, as well as the phylogenetic trees constructed by sector, showed isolates clustering, especially in the animal sector (**Figures 2 and S11**). Based on the distribution of SNPs within the genome-based similarity matrix, a threshold of 40 SNPs was chosen to consider isolates as genetically closely related (**Figure 3A**). All isolates, differing by less than 40 SNPs had the same STc, ST (except two, due to a single mutation in the *fumC* and *gyrB* gene respectively), serotype, *fimH* allele (except one). Five isolates had a different CTX-M variant. A heat map of SNP differences showed intra- and inter-sectoral dissemination of genetically related isolates (**Figure 3B**). A more detailed force-directed layout, representing isolates differing of less than 40 SNPs and harbouring the same ESBL enzyme, showed 169 isolates (38 [46·3%] human, 82 [69·5%] chicken and 49 [63·6%] environmental isolates) gathered in 44 clusters, 15 being mono-sectoral. (**Figure 3C**). The biggest mono-sectoral clusters were observed among chicken isolates, with two (ST716, ST3489) gathering seven to 12 isolates coming from the same, but also from different farms. Three clusters gathered four to five environmental isolates, sampled in different sites (upstream, downstream, and wastewater). Human clusters all had less than four isolates. Twenty-four clusters contained isolates from two sectors (seven human/environment, eight chicken/environment and nine human/chicken). Five clusters (from ST58, ST450, ST602, ST12780 and ST12821) contained 32 (11.6%) isolates from the three sectors. Using the threshold of 10 SNPs, frequently used in the literature to consider *E. coli* isolates as similar,^10^ we constructed another force-directed layout still showing multiple bi- and three tri-sectoral clusters (**Figure 3D**).

**Figure 3.**
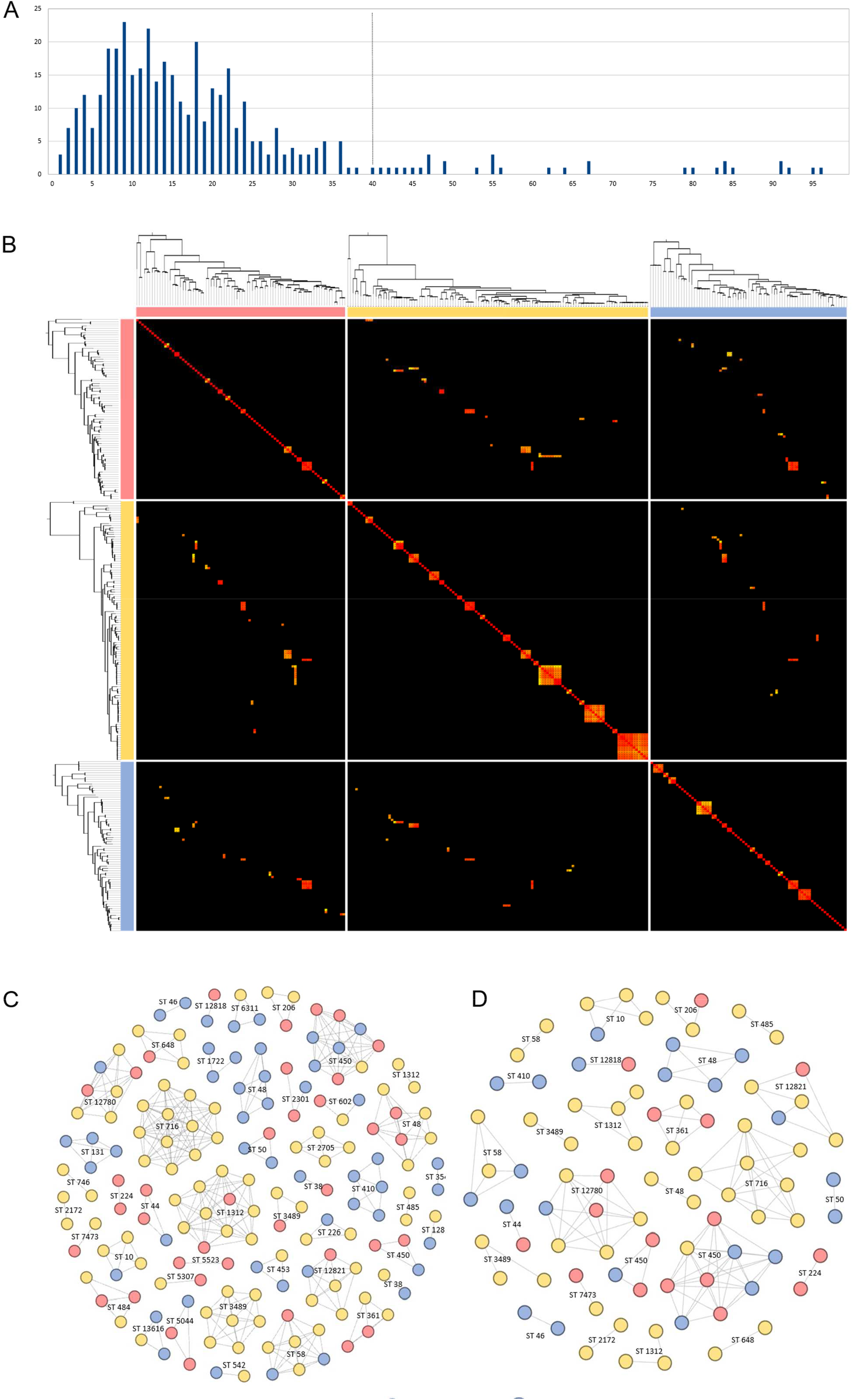
Genetic relatedness of isolates based on single nucleotide polymorphism analysis. **(A)** Distribution of SNPs comparing core genome of ESBL *E. coli* isolates from the three sectors differing of up to 100 SNPs and cut-off chosen (40 SNPs) to consider isolates as genetically related. **(B)** Heat map of SNP differences (0-40 SNPs) between ESBL-*E. coli* isolates grouped by sector (Human – red, Chicken – yellow, Environment – blue). Horizontal and vertical phylogenetic trees are based on core genome sequence per sector. Numbers of SNPs (0-40) in the core genome between different sequenced isolates were interpreted as a distance matrix. Colors in the heat map represent SNP differences as shown in the legend at the right. The diagonal corresponds to the intra-sector comparison with the three main clusters in the chicken sector. Force-directed Fruchterman-Reingold layout representing intra and intersectoral dissemination of isolates differing by **(C)** less than 40 SNPs and (D) up to 10 SNPs. Branch lengths are not representative of SNP distances.

### Plasmid circulation

We secondly investigated intra- and inter-sectoral circulation of plasmids carrying ESBL genes. As a limited number of isolates harboured other plasmid types, we focused the analysis on IncF and IncY plasmids.

#### ESBL-carrying IncF plasmids

ESBL-IncF plasmids were distributed into 15 different pMLST subtypes, the primary being [F31/36:A4:B1] (20·6%, n=13/63), followed by [F-:A-:B53] and [F1:A1:B49] (12·7%, n=8/63 each). IncF plasmids with undetermined pMLST [F-:A-:B-], were the most frequent (22%, n=14/63) but were distributed in two distinct groups, including a highly conserved phage plasmid carrying *bla*_CTX-M-55_. All plasmid subtypes containing more than one isolate (53%, n=8/15) were observed in at least two different sectors, carried by *E. coli* isolates with different genetic backgrounds. The [F31/36:A4:B1] (99% sequence homology) and [F-:A-:B53] (95% sequence homology) subtypes, were harboured respectively by seven and six genetically different *E. coli* isolates and observed in all sectors (**Figure 4A**).

**Figure 4.**
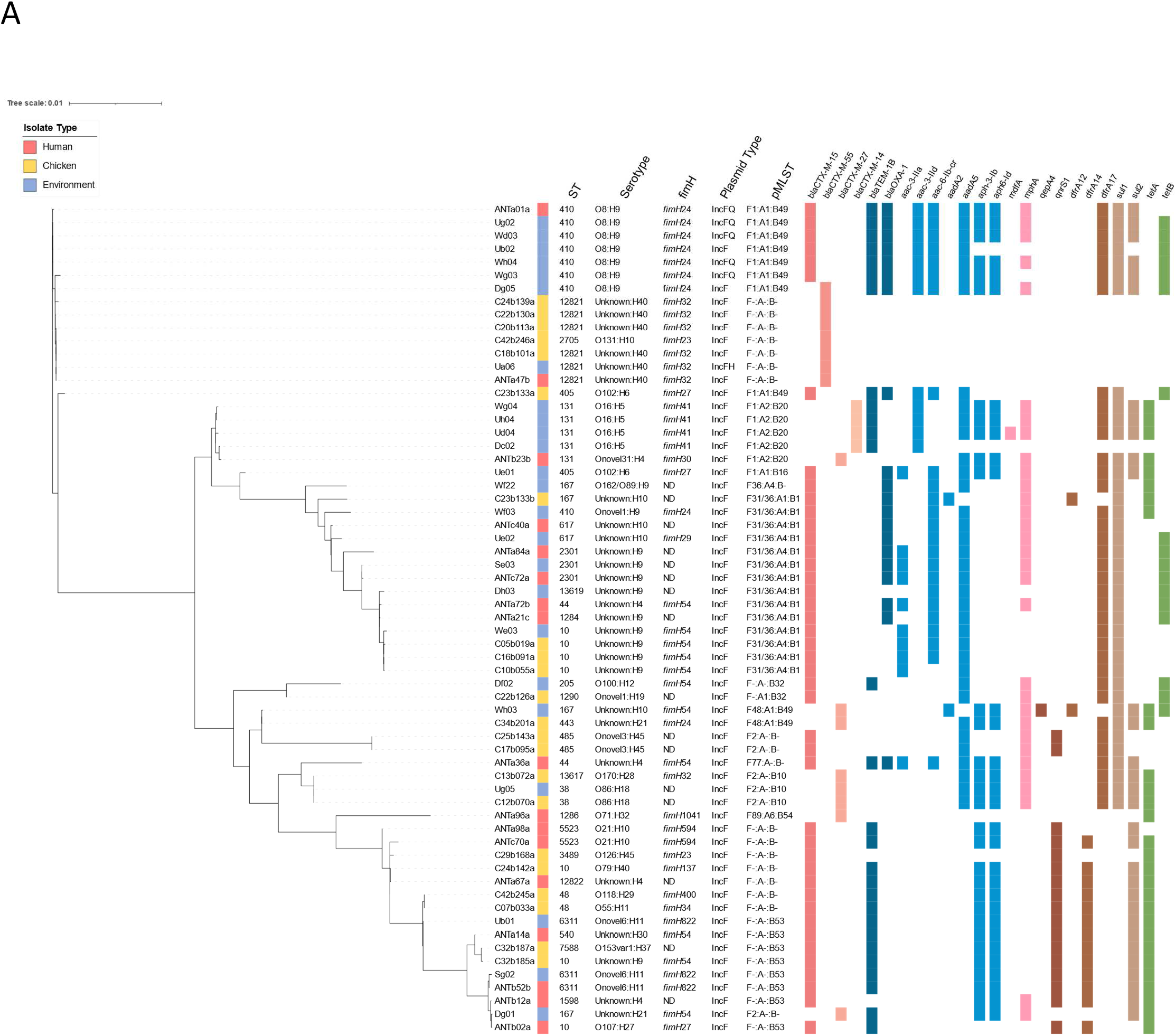
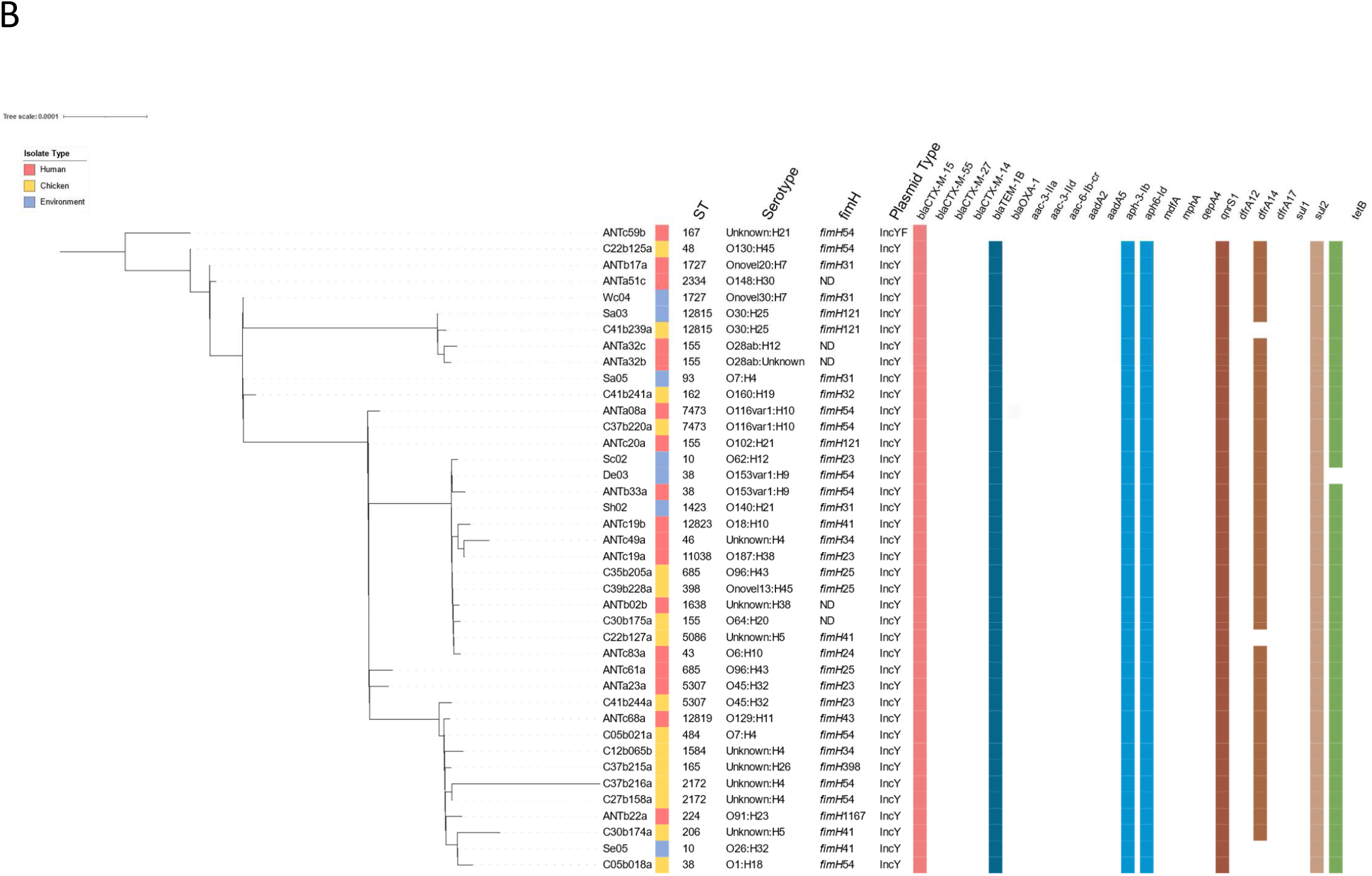
Pangenome trees of IncF and IncY plasmids. The trees were constructed from the presence/absence matrix of genes. Strip one and two contain the name of the isolate and its sector origin. Strips three to five represent isolates ST, serotype and FimH allele. Strips six and seven represent plasmid type and plasmid MLST type (for IncF plasmids only). Coloured strips represent plasmid-borne resistance genes. Scales differ between the two trees. **(A)** Pangenome tree of IncF plasmids. **(B)** Pangenome tree of IncY plasmids. The IncY/F plasmid was included in the IncY tree based on their structural similarity (*vir* transfer system). Two isolates were excluded from the alignment due to insufficient quality of the sequencing.

#### ESBL- carrying IncY plasmids

The ESBL-IncY plasmids showed >99% sequence homology (with 70-100% coverage) and a highly conserved resistance gene content (**Figure 4B**). With the exception of *tet(A)* and *dfrA14*, lost by deletion in two and five plasmids respectively, all other ESBL-IncY plasmids carried *qnrS1, bla*_CTX-M-15_, *bla*_TEM-1B_, *aph(6)-Id, aph(3”)-Ib, sul2* and *dfrA14* genes and two copies of *tet(A)*. Alignment of the IncY genomes confirmed this homology and highlighted the presence of two common unstable zones, one starting with an IS26 and containing the *xerC* (tyrosine recombinase) and *betU* genes and the other starting with an IS1R and affecting more or less the *virB* gene cluster involved in conjugation (**Figure S12**). ESBL-IncY plasmids were observed in all three sectors within 34 *E. coli* isolates with different genetic backgrounds.

Finally, to visualize links among plasmids, strains, and sectors, we constructed a Sankey diagram, which confirmed the significant intra- and inter-sectoral circulation of both ESBL-*E. coli* strains and ESBL-plasmids (**Figure 5**).

**Figure 5.**
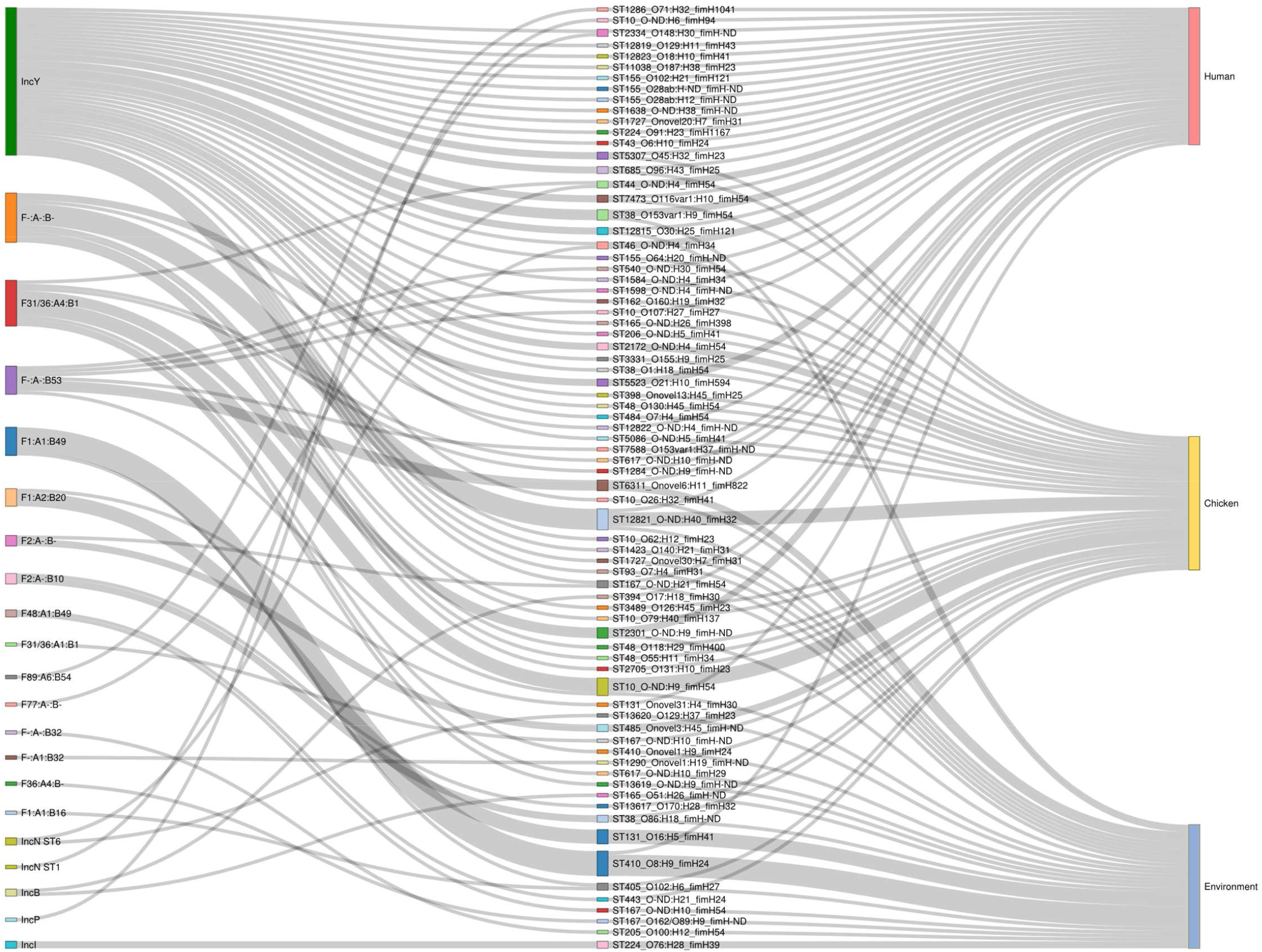
Sankey diagram representing relationships between ESBL carrying plasmids (left), ESBL-*E. coli* isolates (middle) and sampling sectors (right). The grey lines are connecting the three items. For easier reading, in this diagram, isolates were gathered when having the same ST, serotype and *fimH* allele.

## Discussion

To our knowledge, the present study is the first to address the circulation of antibiotic resistance from a One Health perspective in a low-income country, considering both ESBL-*E. coli* strains and ESBL-carrying plasmids in humans, farm chickens and the environment. It is also the first to have carried out genomic analysis of the results of the implementation of the WHO Tricycle protocol. Thanks to high-quality sampling, performed on the same temporal scale and in a limited geographical area, we were able to evidence significant circulation of ESBL-*E. coli* strains and ESBL-plasmids within and among all three sectors despite populations of differing genetic structure.

In our study ESBL-*E. coli* prevalence in healthy pregnant women was nearly 30%, which is an increase from previous studies, reporting a prevalence of ESBL producing *Enterobacterales* in the Malagasy community of 10% in 2009 ^11^ and 19% in 2015.^12^ Concerning the animal sector, the ESBL-*E. coli* prevalence in chickens reached 57%. ESBL-*E. coli* was detected in all farms, unlike a study conducted in Madagascar in 2016, in which ESBL producing *Enterobacterales* were found in 70% of participating farms.^13^ Finally, all environmental samples contained ESBL-*E. coli*, with increasing concentrations from upstream and downstream surface water to wastewater and slaughterhouse effluents, underscoring the impact of dense human populations and of untreated wastewater on ESBL-*E. coli* spread in the environment.

The only risk factor identified for human and animal ESBL-*E. coli* carriage was the wet season during which temperatures and rainfall are higher than in the dry season in the Antananarivo area. We have already reported a correlation between ESBL-*E. coli* prevalence carriage in humans and the wet season in a previous study in Madagascar, as well as other studies.^7,14,15^ In contrast, we did not find any publications reporting a positive correlation between seasonality and ESBL-*E. coli* carriage in farm animals. The rainy period is well known for favouring faecal-oral transmission of enteric bacteria.^16^ *In vitro* models also suggest that growth rates and horizontal gene transfer may be enhanced by higher temperatures and humidity,^17,18^ making global warming an additional threat to the dissemination of antibiotic resistance.

In all isolates, the ESBL phenotype was due to a *bla*_CTX-M_ gene, *bla*_CTX-M-15_ being the most frequent whatever the sector, which is consistent with the global ESBL epidemiology. Surprisingly, we observed chromosomal integration of *bla*_CTX-M_ genes in more than half (57%) of the isolates. High rates of *bla*_CTX-M_ chromosomal integration are being increasingly reported, reaching, for example, 64% in a Japanese study.^19^ Moreover, we detected multiple chromosomal insertion sites over a large part of the genome suggesting frequent and repeated events. Of note, most of the insertions were localized in regions outside the *oriC* macrodomain and encompassing the terminus of replication, a region known for hyper-recombination.^20^ The chromosomal integration is considered to provide stabilization of *bla*_CTX-M_ genes at a lower fitness cost in comparison to a plasmid. This enables vertical transmission and *bla*_CTX-M_ persistence even in the absence of selective pressure.^21^ However, its impact on the *bla*_CTX-M_ dissemination in the future, is hardly predictable.

As already described in the literature,^5^ WGS analysis confirmed different population structures within the three sectors with variable gene contents (virulence, resistance, replicons), STs, genome sizes, as well as variation in diversity. Despite these ecological peculiarities, we observed an intersectoral circulation of strains. We performed a SNP analysis of strains differing of less than 40 SNPs, a threshold based on SNPs distribution in the entire collection. This analysis revealed multiple clusters of closely related isolates, gathering isolates from the same sector (especially chickens) but also from two and from all three sectors, reinforcing the hypothesis of intersectoral transmission. SNP cut-offs for considering genetically related isolates are highly variable in the literature, ranging from 10 to 100.^22,23^ Therefore, we performed a more stringent analysis, retaining only isolates differing of up to 10 SNPs, the threshold frequently used to consider *E. coli* isolates as similar,^23^ and we still observed clusters of closely related isolates belonging to the three sectors, evidencing intersectoral ESBL-*E. coli* dissemination in the Antananarivo area. Most studies analysing the intersectoral circulation of ESBL-*E. coli* have identified rather genetically distinct populations of human, animal and environmental isolates, with inter-human transmission being the major source for community colonization.^24–27^ However, these studies have rarely investigated isolates collected at the same temporo-spatial scale.^24,25,27^ Moreover, they included human strains, isolated from invasive infections, and were mainly conducted in high-income countries, where *E. coli* strains from B2 and D phylogroups have progressively outpaced commensal phylogroups A and B1.^5,27^ The discrepancy with our results could be explained by different behaviours in developing countries driven by local socio-economic factors, availability of over-the-counter antimicrobials, or insufficient sanitation. This hypothesis is supported by a Ghanaian study also showing that closely related strains were shared by humans and animals.^22^

Additionally, we investigated the intra- and inter-sectoral circulation of ESBL carrying plasmids. Predominant ESBL-plasmids were of IncF type, well known for their high dissemination ability, especially F31/36:A4:B1 and F-:A-:B53, which we identified in *E. coli* of different genetic backgrounds and from different sectors.^28,29^ In a previous study we described an unusual prevalence of a highly conserved ESBL-IncY plasmid in *E. coli* isolates from pregnant Malagasy women.^7^ Here, we confirmed the significant dissemination of the same ESBL-IncY plasmid also in chickens and in the environment. IncY plasmids are usually considered as phage-plasmids, but the IncY described here do not have genes coding for phage proteins and contain a *virB* conjugation system.^30^ The detection of the same IncY plasmid and some IncF subtypes in the three sectors, harboured by *E. coli* of various genetic background, showed another pathway of dissemination of ESBL resistance in a One Health context in the Antananarivo area.

The present study has several limitations. First, ESBL-*E. coli* strains isolated from blood cultures representing the human hospital component of Tricycle could not be included. Due to their cost, blood cultures are rarely prescribed in Madagascar and the rare ESBL-*E. coli* isolates identified were not stored. Second, the Tricycle protocol requires three to five colonies per sample, and despite this, these are still the predominant isolates within a sample and may not represent the total genetic diversity of ESBL-*E. coli* carried. Finally, although our results showed circulation of ESBL-*E. coli* and ESBL-plasmids between sectors, the direction of the transmission routes could not be determined.

## Conclusion

By implementing the One Health Tricycle surveillance project in Antananarivo (Madagascar), we detected a high ESBL-*E. coli* prevalence in healthy pregnant women, farm chickens and the environment. Thanks to WGS analysis, we observed the circulation among all three sectors of both ESBL-*E. coli* strains and ESBL-carrying plasmids, despite different sectoral population structures. In order to tackle antibiotic resistance, measures on global antibiotic consumption need to be accompanied by actions to limit the transmission of resistance between the three sectors.

## Data Availability

All data produced in the present study are available upon reasonable request to the authors.

https://www.ebi.ac.uk/ena/browser/view/PRJEB56633

## Data Availability Statement

The original contributions presented in the study are publicly available and can be found at: https://www.ebi.ac.uk/ena/browser/home (accession number PRJEB56633).

## Acknowledgments

We would like to thank the Oxford Genomics Centre at the Wellcome Centre for Human Genetics (funded by Wellcome Trust, grant reference 203141/Z/16/Z), for the generation and initial processing of the sequencing data).

## Supplementary methods

### Study design and setting

Sampling was performed in Antananarivo, the capital of Madagascar with a population of 3· 2 million of inhabitants.

Healthy pregnant women were enrolled, from July 2018 to April 2019, after signing a written informed consent. Rectal swabs were collected in three maternity wards in Antananarivo (Joseph Raseta Befelatanana hospital [JRB] - July to August 2018, Mère-Enfant Tsaralalana hospital [TSA] - July to October 2018 and Joseph Ravoahangy Andrianavalona hospital [JRA] - March to April 2019) and sent in each hospital laboratory.

Alive chickens were purchased, from April 2018 to March 2019, in batches of six chickens per week, in 27 different wet markets in Antananarivo (41 purchasing campaigns). Chickens from the same batch came from the same farm and were placed in the same cage during transport.

Environmental sampling was conducted, from April 2018 to April 2019, on surface water from Ikopa river upstream (5· 5km from downtown) and downstream (5· 4km from downtown), from a wastewater channel (in the city center) and from a slaughterhouse sewage (5· 4km from downtown) (7 sampling campaigns) (Figure S1).

Animal and environmental samples were processed at the CICM (Charles Merieux Center of Infectious Disease), Antananarivo.

### Microbiology

Microbiological methods were performed according to the Tricycle protocol recommendations in Antananarivo.^1^ All indole positive isolates, suspect of ESBL production, were sent to Merieux Fondation (Lyon, France) for verification of identification (Vitek-2, bioMérieux, Marcy L’Étoile, France) and determination of antimicrobial susceptibility by the disc diffusion method (Mueller-Hinton agar, Sigma-Aldrich, St. Louis, USA) according to EUCAST recommendations (www.eucast.org).

Antimicrobial susceptibility was determined using a large panel of antibiotic disks: amoxicillin, amoxicillin/clavulanic acid, ticarcillin, ticarcillin/clavulanic acid, piperacillin, piperacillin/tazobactam, cefotaxime, cefalexin, ceftazidime, cefepime, cefoxitin, aztreonam, temocillin, mecillinam, imipenem, meropenem, ertapenem, fosfomycin, colistin, chloramphenicol, nalidixic acid, ofloxacin, ciprofloxacin, levofloxacin, amikacin, gentamicin, netilmicin, tobramycin, tetracycline, sulfonamides, trimethoprim, and trimethoprim/sulfamethoxazole.

For each sample, all isolates with distinct antibiotic profiles, based on the susceptibility and diameters of the 32 antibiotics on the antibiogram, were selected for whole genome sequencing by the Illumina technology.

### Illumina sequencing

At Oxford Genomics Centre, libraries were prepared with the Celero DNA-Seq PCR free kit (NuGen Technologies, Redwood, CA, United States) for library preparation and sequencing was performed on NextSeq 500 and NovaSeq 6000 platforms (Illumina, San Diego, CA, United States).

Reads quality was first checked using FastQC v0.11.8. Reads were trimmed by Trim Galore v0.4.4 (quality > 30, length > 50bp).

Contigs were assembled with SPAdes v3.11.1 and contigs longer than 500bp were further analyzed. Phylogroups were determined using the standalone ClermonTyping software.^2^ Serotypes and Sequence types (ST) were determined using SerotypeFinder v.1.0.0^3^ and MLST Finder v1.8^4^ (Warwick University ST scheme) respectively. Sequence type complexes (STc) were retrieved from Enterobase (https://enterobase.warwick.ac.uk/species/index/ecoli).

Resistance genes and plasmid replicons were identified using AMRFinder v2019.04.29 and ResFinder v2021.09.23^5^ and PlasmidFinder v2020-07-13^6^ respectively with DIAMOND v0.9.22.123 software.^7^ Virulence genes were detected using Abricate software v.0.9.8^8^ to query Virulence Finder v. 2016-02-18^9^ and VfDB v. 2018-11-05^10^ databases and an in house developed virulence factors database.^8^

Roary software v3.12^11^ and IQTree v1.6.9^12^ were used to construct a maximum likelihood phylogenetic tree (GTR model, 1000 bootstrap) with *Escherichia fergusonii* complete genome [strain ATCC35471, GenBank accession: GCA_012811895.1] as the root. Graphical representation was generated using iTOL v6.3.^13^ SNPs were calculated by Parsnp v1.12.^14^ Comparison of isolates were performed using two cut-offs. The first was chosen to consider isolates as closely related, depending on the distribution of the SNPs in the studied population. A second cut-off of 10 SNPs was chosen, according to the literature to consider *E. coli* isolates as similar.^15^

For *bla*_CTX-M_ genes located on the chromosome, sequences upstream and downstream of *bla*_CTX-M_ were identified using Geneious software v5.1.7 (https://www.geneious.com). Insertion sites were set on the *E. coli* K-12 linkage map.^16^

### Oxford Nanopore Sequencing

Libraries were prepared using SQK-LSK109, EXP-NBD104, EXP-NBD114 kits (Oxford Nanopore Technologies, Oxford, UK). WGS was performed on a MinION platform (Oxford Nanopore Technologies,)

Plasmid sequences were reconstructed using Illumina and Nanopore reads with Unicycler v0.4.9b software.^17^ Resistance genes and plasmid replicons were identified using ResFinder v.2020-10-20^5^ and PlasmidFinder v.2020-07-13^6^ respectively with DIAMOND v0.9.22.123 software.^7^

All plasmid sequences were compared using BLAST v2.9.0. Roary software v3.12^11^ was used to define the plasmid pangenome and to generate an alignment file with a matrix based on the presence/absence of genes. IQTree v1.6.9^12^ was used to calculate the phylogenetic tree using the alignment of the pangenome. Graphical representation was generated using iTOL v6.3^13^. Annotation was performed using PROKKA v1.14.6 ^18^. Graphic presentation was generated with BRIG v0.95.^19^

### Statistical analysis

Epidemiological data collected were (i) for pregnant women: age, education level, access to electricity, water and toilets, presence of pet/farm animals and recent antibiotics intake (in the previous three months), (ii) for chickens: weight and (iii) for all, rainfall and mean temperatures at the time of sampling, retrieved from https://www.historique-meteo.net/afrique/madagascar/antananarivo/.

We compared the proportion of each gene from different sectors by Fisher exact test. The reported p-value was adjusted by method Benjamini-Hochberg. Descriptive statistics included counts and percentages for the description of binary variables and mean and interquartile range for quantitative variables. Factors associated with ESBL-*E. coli* carriage were identified with univariable logistic regressions with carriage as dependent variable. We reported odds-ratio estimates with 95% profile confidence intervals and p-values from the likelihood ratio test.

The threshold for statistical significance was 0·05 and all analyses were performed with R software v4.1.1.

To evaluate the genetic diversity among strains within each sector (human, chicken, environment) we computed the pairwise nucleotide diversity (π)^20^ from the alignment obtained with Roary software v3.12.^11^ We also computed the intra-sectoral genetic diversity (human strains versus human strains, chicken strains versus chicken strains and environment strains versus environment strains) and the inter-sectoral genetic diversity (human strains versus chicken strains, human strains versus environment strains and chicken strains versus environment strains). We used t tests to compare the genetic diversity and the Benjamini & Hochberg correction for multiple tests when appropriate.

**Figure S1.**
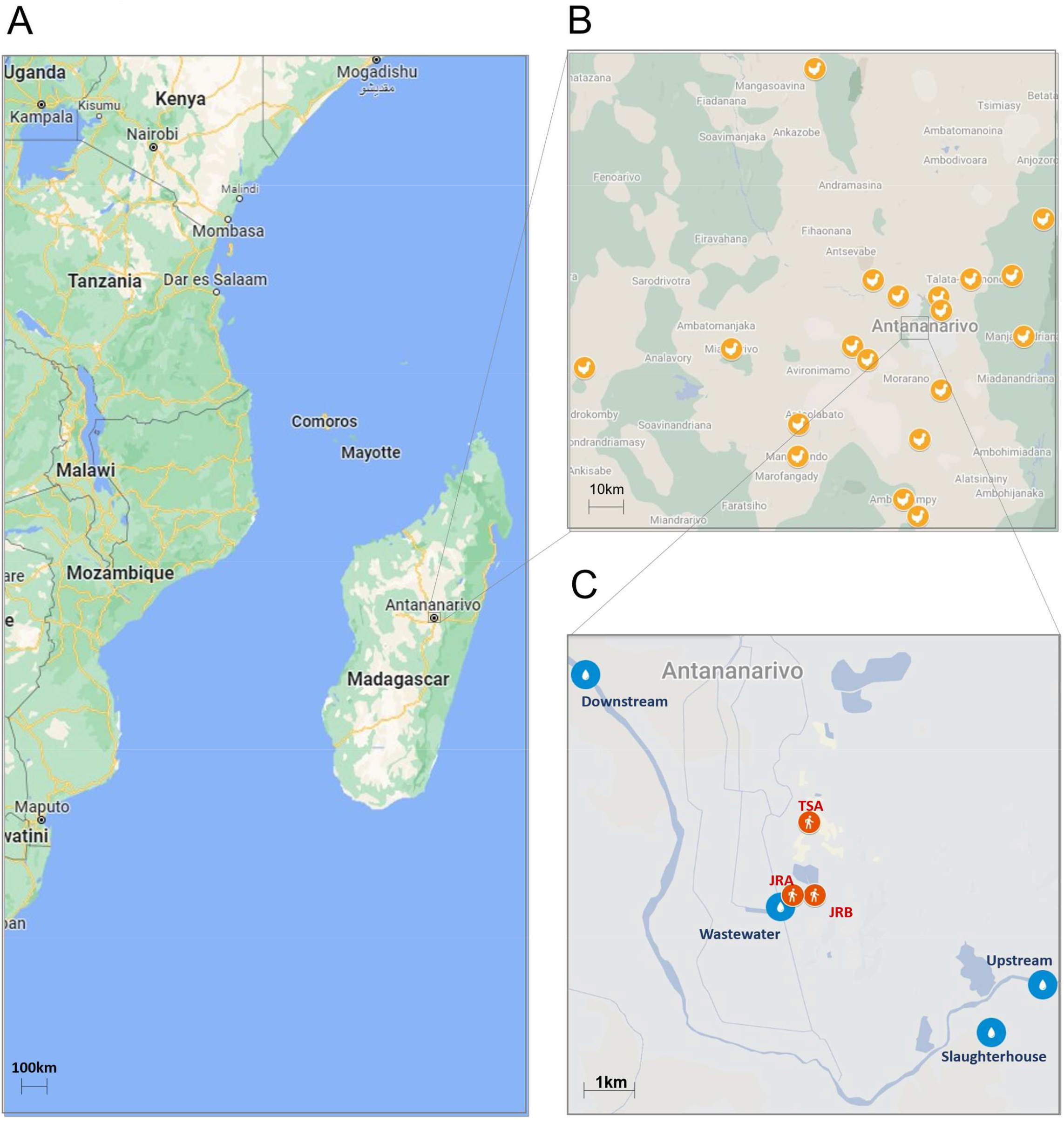
Sampling locations. (A) Location of Madagascar and Antananarivo. (B) Animal sector sampling locations. Animal sector farming locations are represented by yellow pins. (C) Environmental and human sector sampling locations. Blue pins indicate the environmental sampling sites. Red pins indicate the three maternity wards, Joseph Raseta Befelatanana hospital (JRB), Mère-Enfant Tsaralalana hospital (TSA), and Joseph Ravoahangy Andrianavalona hospital (JRA).

**Table S1.**
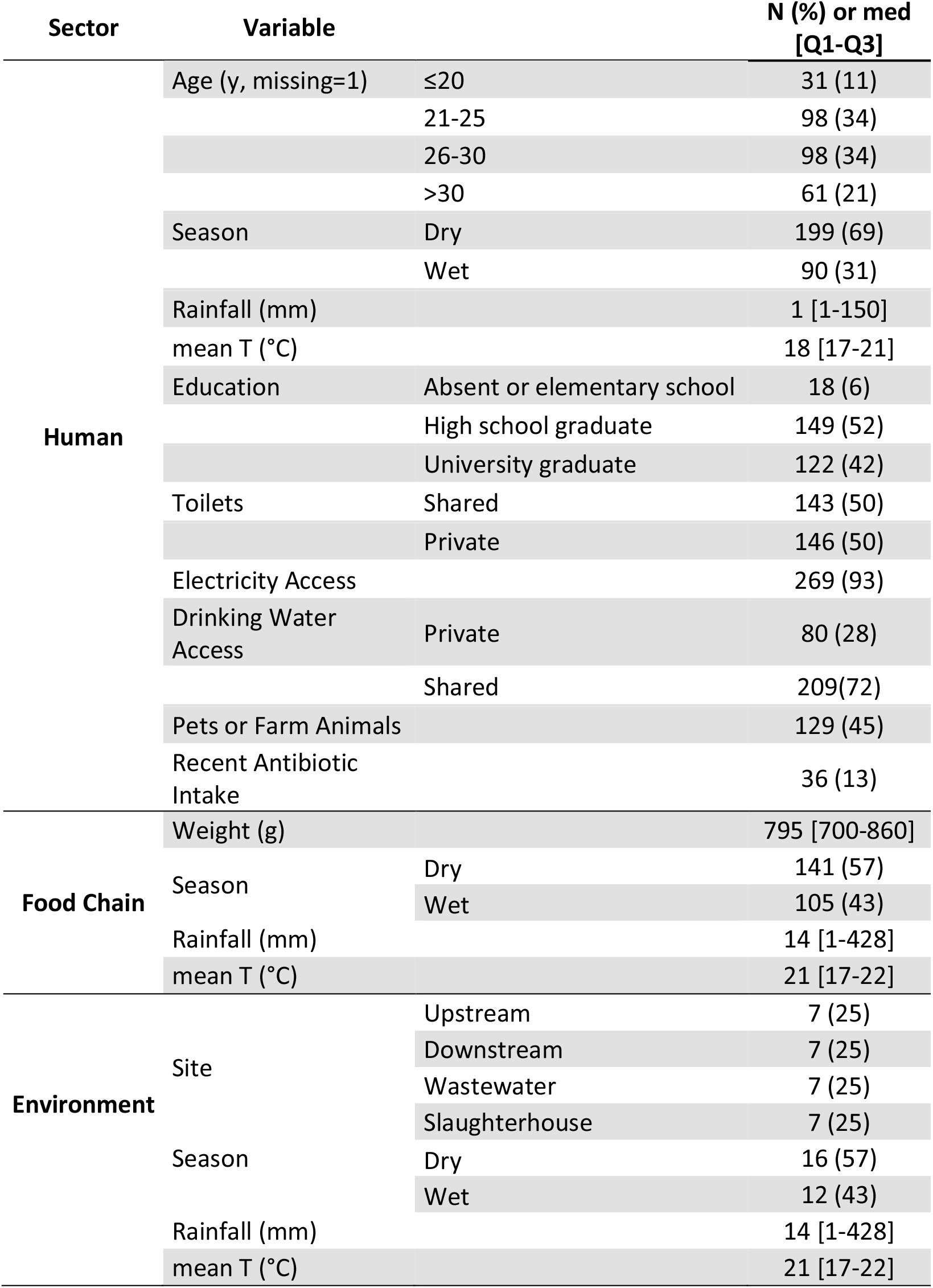
Characteristics of human, chicken and environmental samples and of weather conditions during sampling in Antananarivo, Madagascar, July 2018 to April 2019.

**Figure S2.**
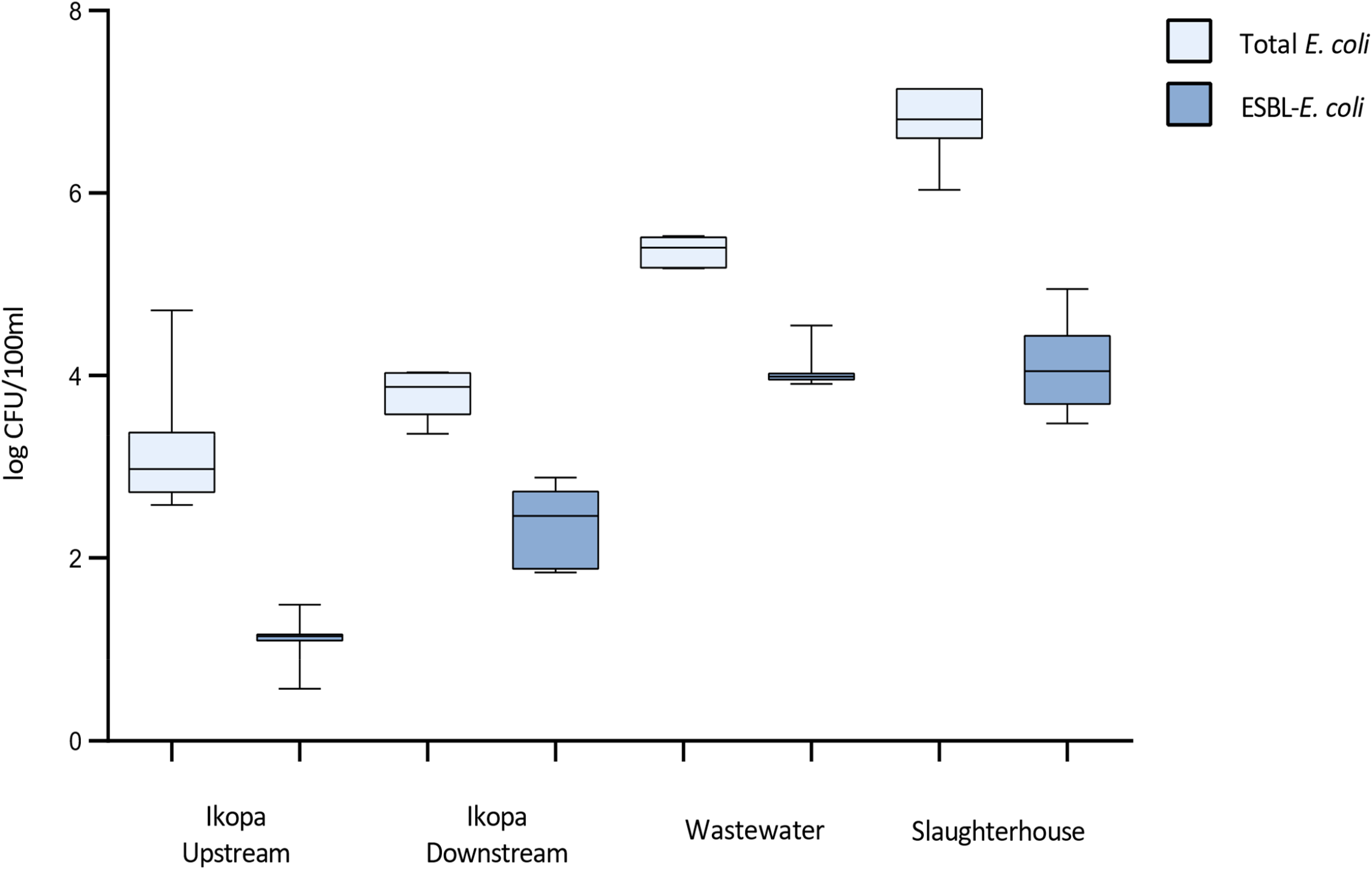
Mean *E. coli* and ESBL-*E. coli* concentrations in Ikopa upstream and downstream surface water, wastewater and slaughterhouse effluents, expressed in logCFU/100ml.

**Figure S3.**
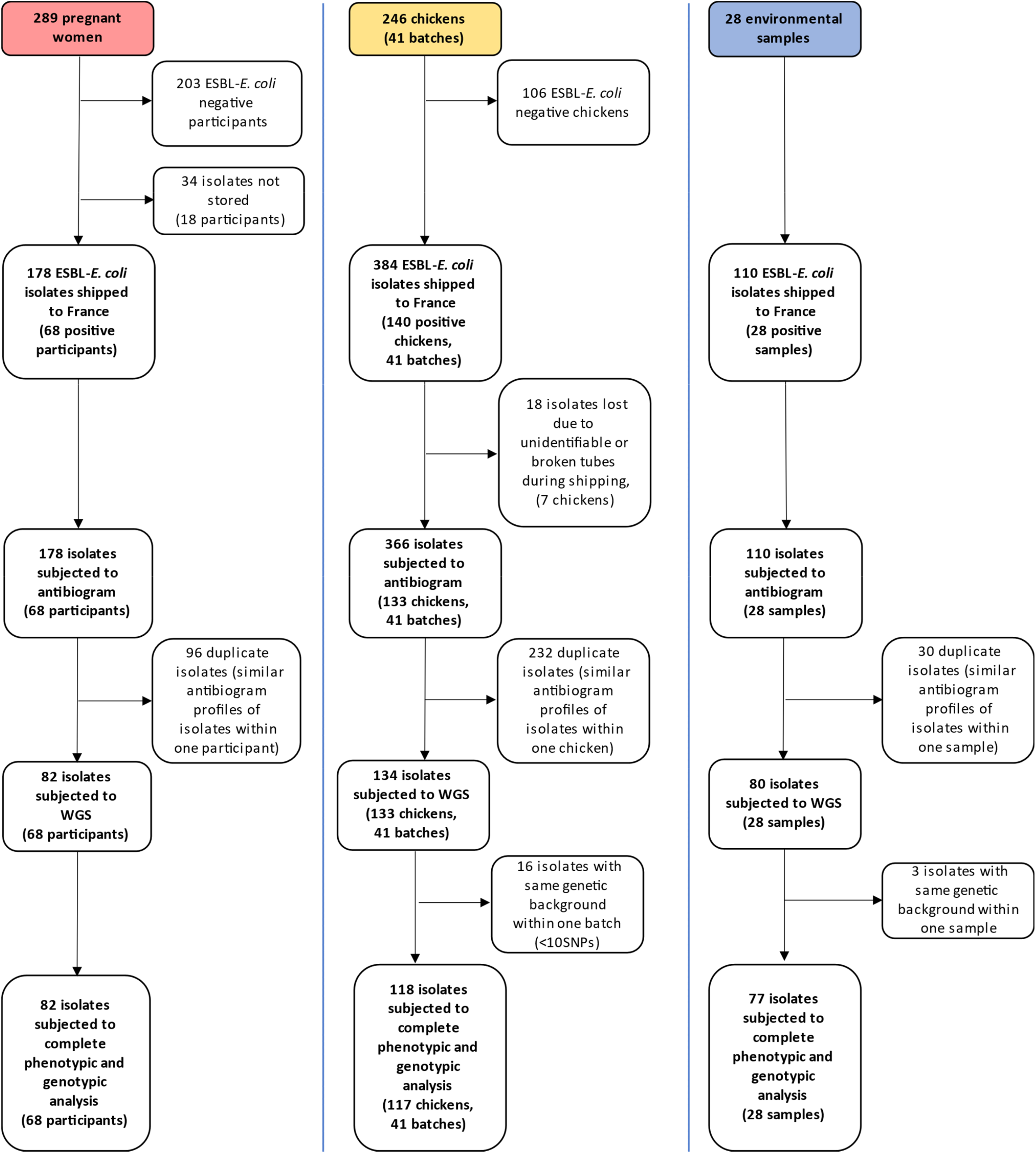
Flow chart representing the evolution of the number of isolates along study progress.

**Figure S4.**
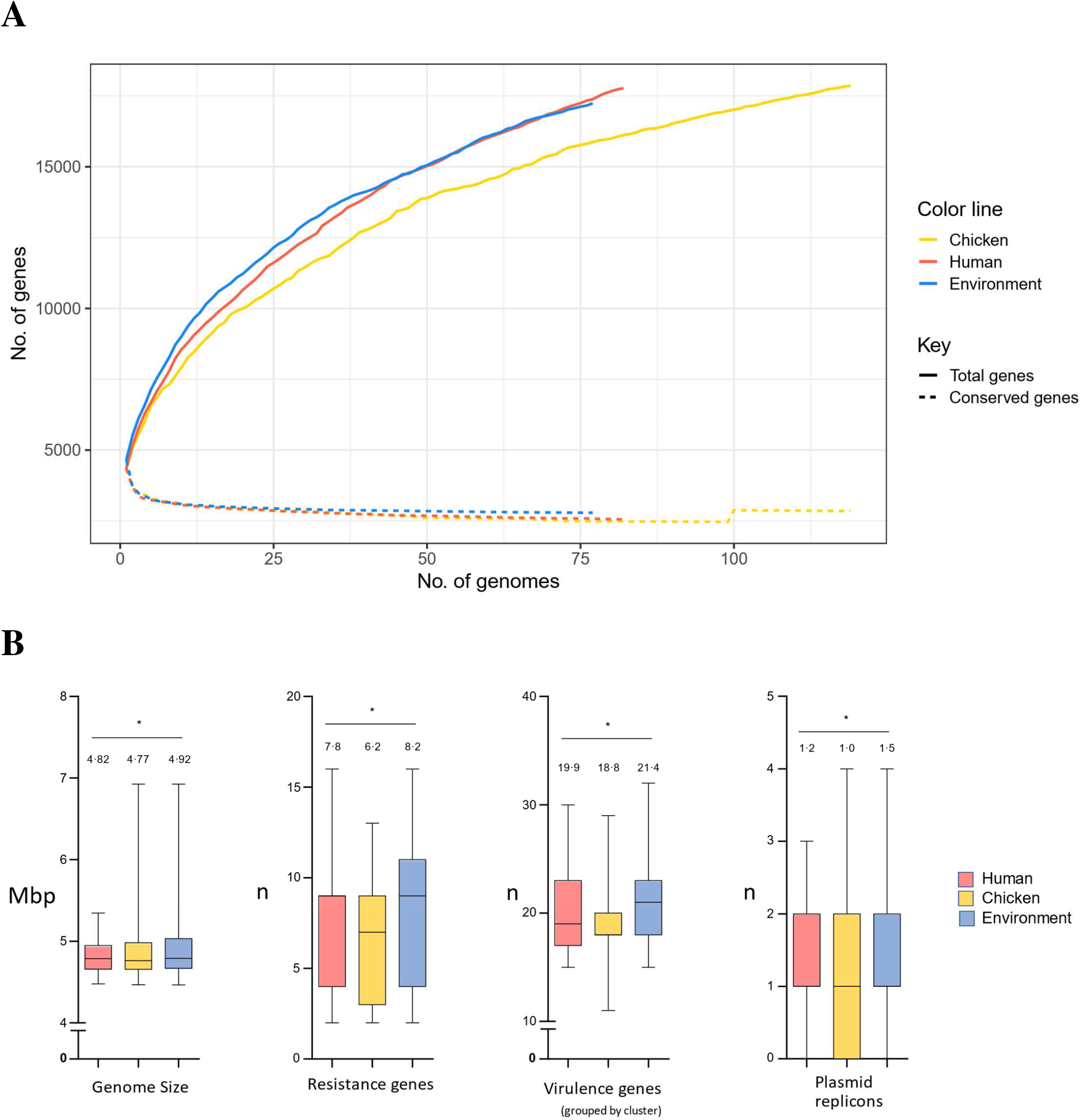
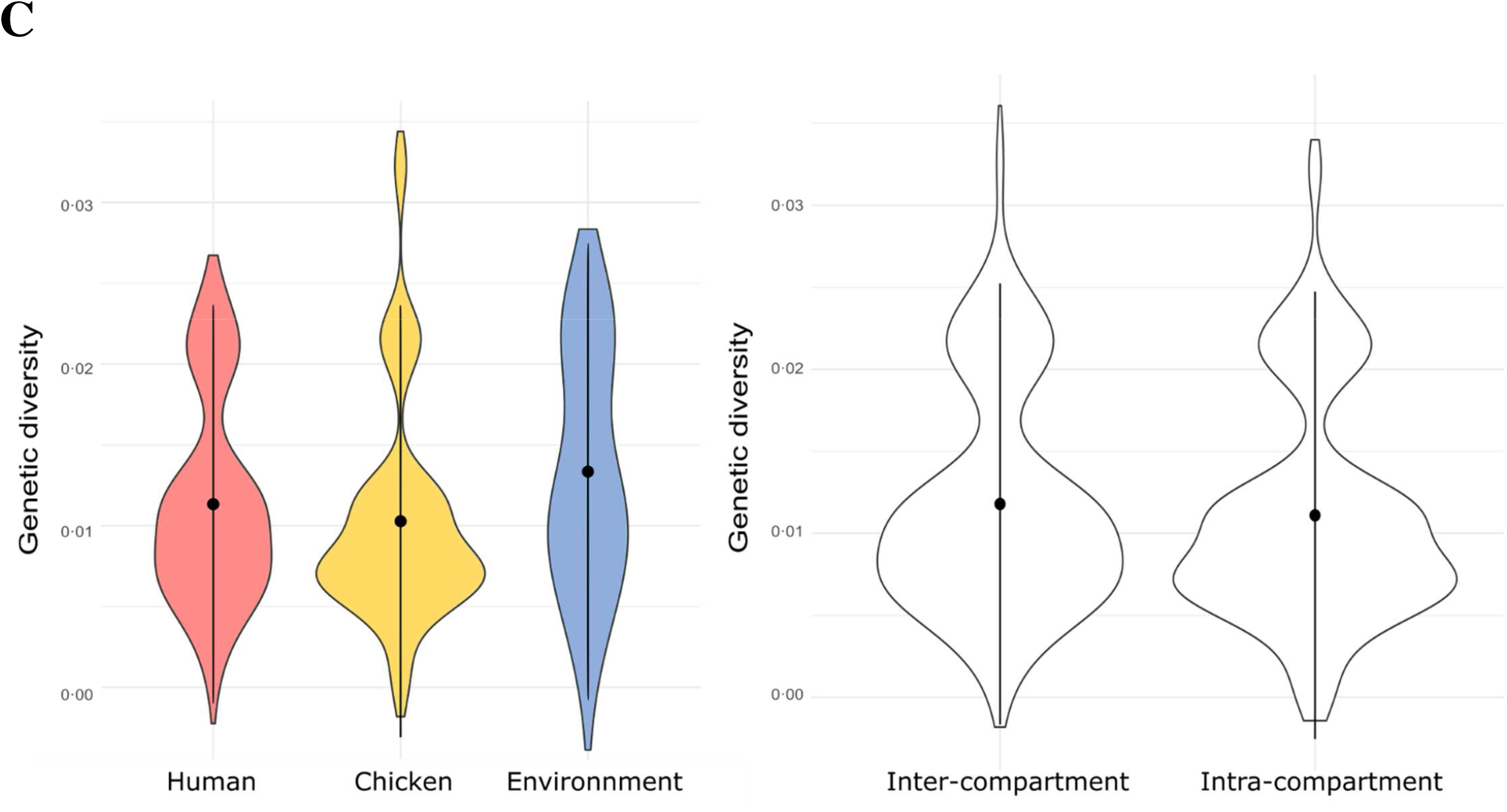
Representation of gene content of isolates from the three sectors. (A) Conserved core and pangenome gene content (B) Mean number of resistance genes, virulence genes (one per gene cluster/operon), plasmid replicons, and mean genome size (Mbp) per isolate (C) Intra- and inter-sector genetic diversity. Asterisks indicate significant differences between sectors.

**Figure S5.**
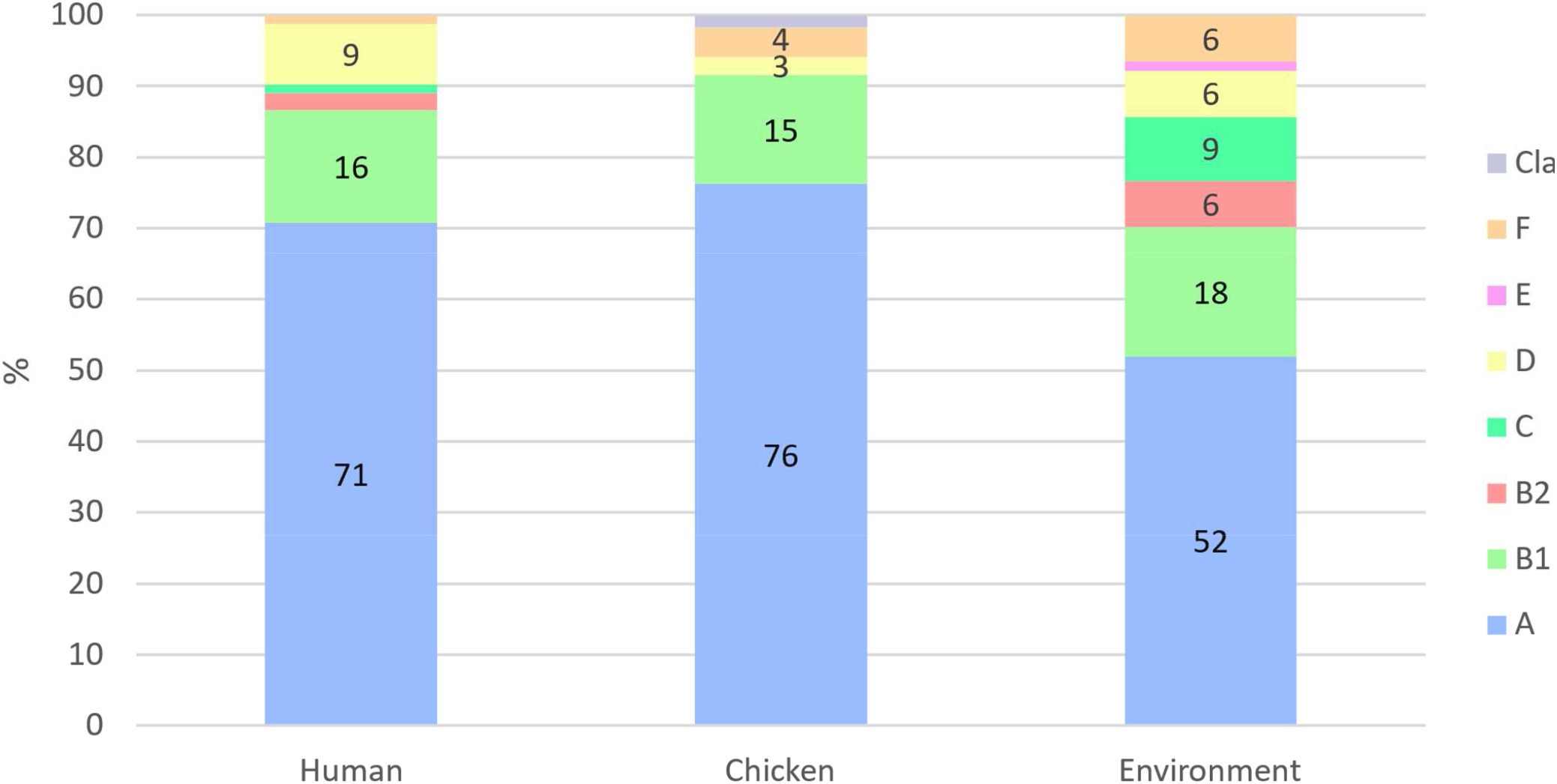
Distribution of *E. coli* phylogenetic groups among the three sectors.

**Figure S6.**
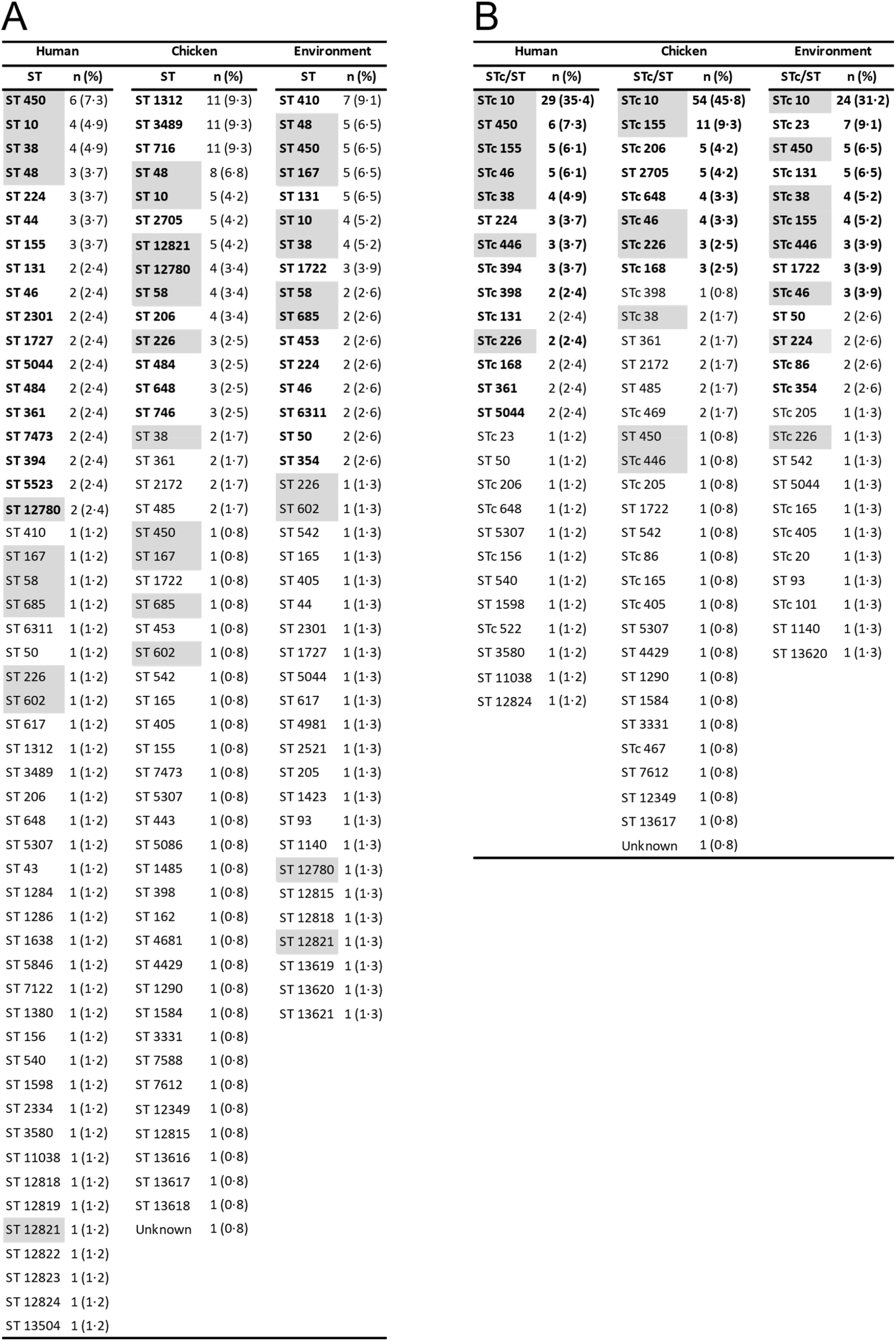
Distribution of sequence types (ST) and clonal complexes (STc) among the three sectors. (A) Distribution of ST. (B) Distribution of STc in the three sectors (when no STc was available, the ST was noted). STs and STc in bold represent the most frequently (> 2%) observed in each sector. Cells highlighted in grey indicate ST or STc recovered from all three sectors.

**Figure S7.**
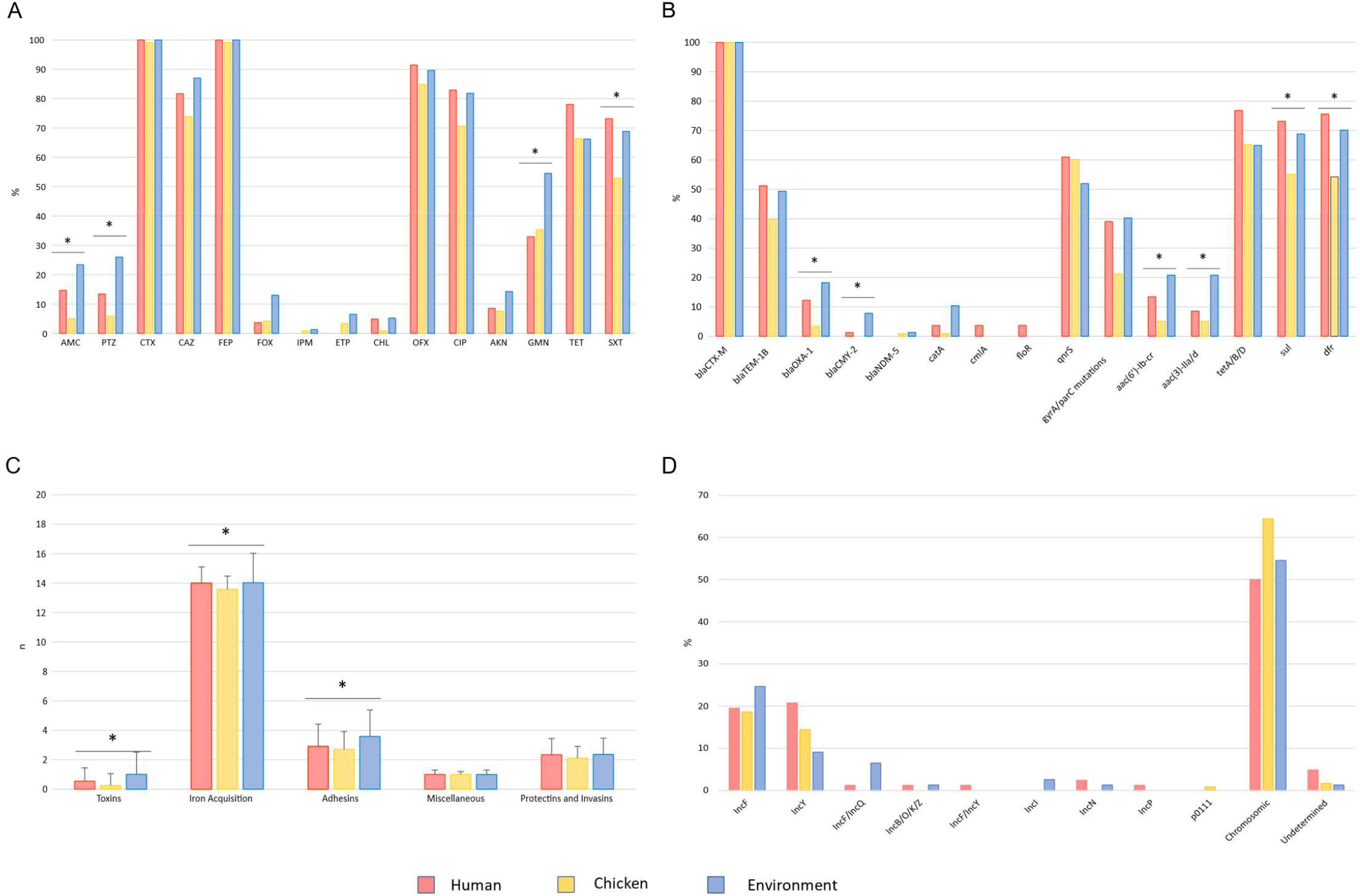
Prevalence of phenotypic antibiotic resistance, resistance and virulence gene content, and *bla*_CTX-M_ location. (A) Prevalence of phenotypic resistance to main antibiotics. Percentages correspond to resistant and intermediately resistant isolates for each antibiotic. (B) Prevalence of the main resistance genes. (C) Mean number of virulence genes per isolate grouped by category (toxins, adhesins, iron acquisition, protectins and invasins, miscellaneous). (D) Prevalence of *bla*_CTX-M_ location, chromosomic or plasmidic with type of plasmid. Asterisks indicate significant differences between sectors.

**Figure S8.**
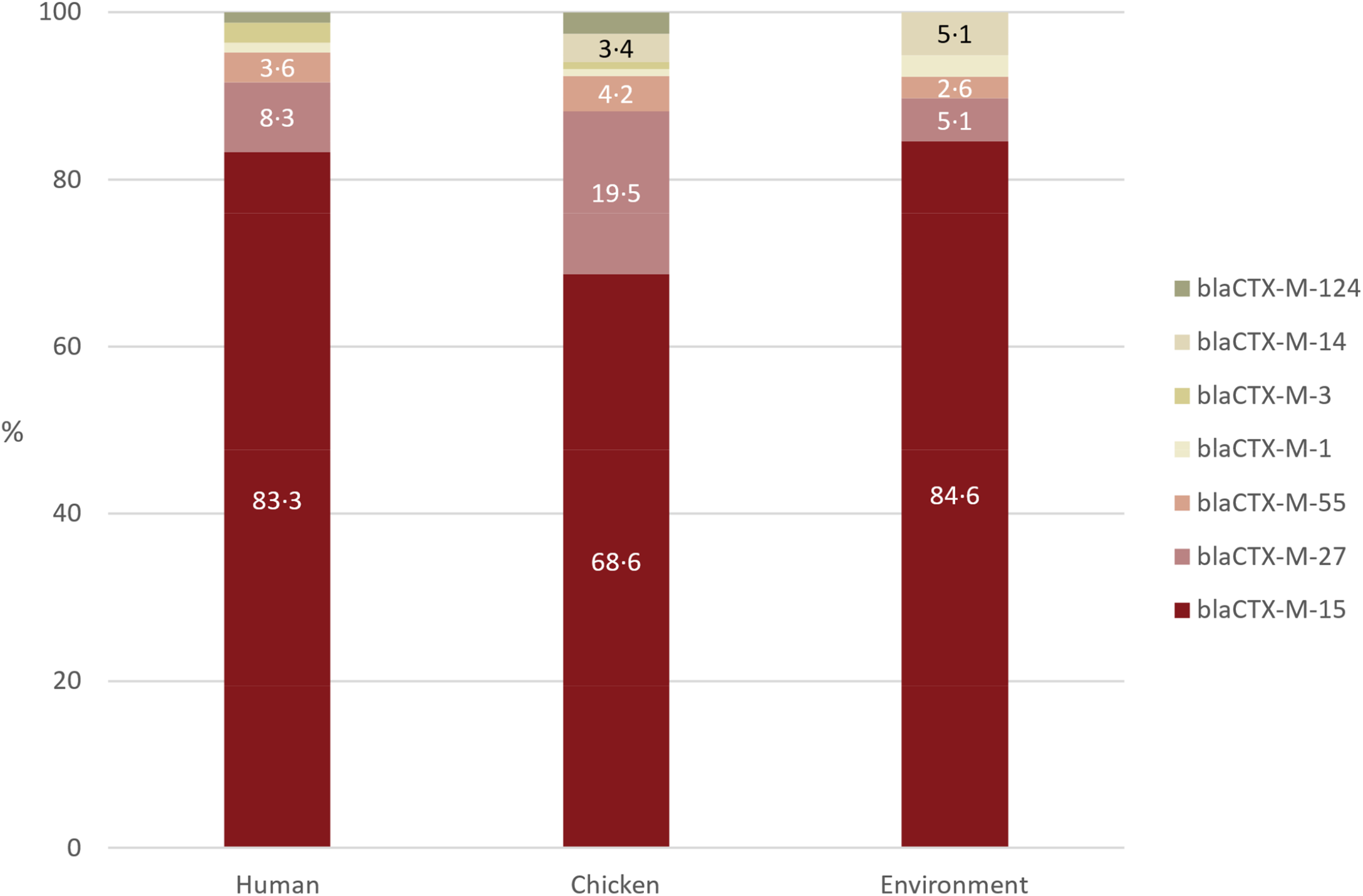
Distribution of *bla*_CTX-M_ genes among the three sectors.

**Figure S9.**
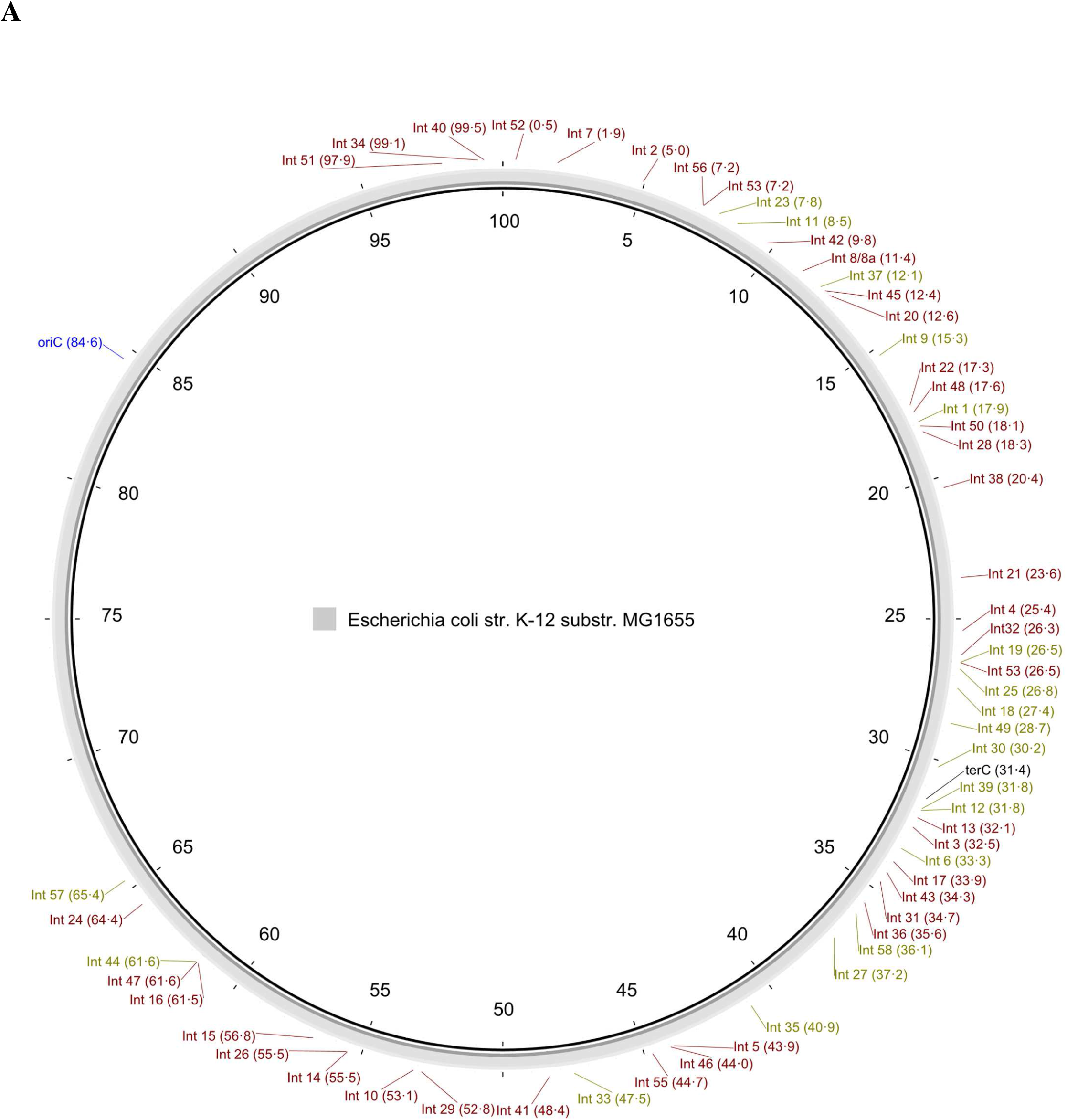
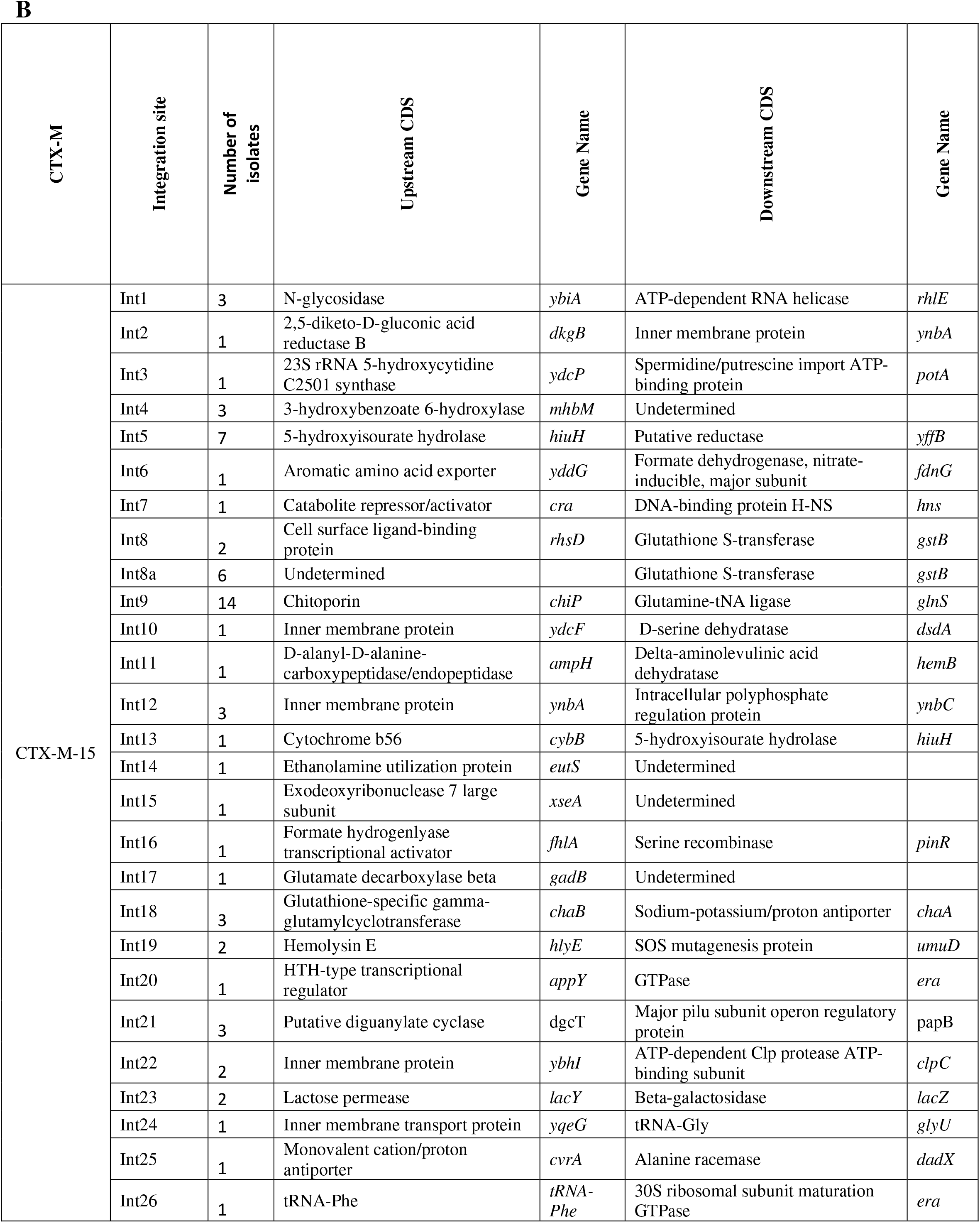
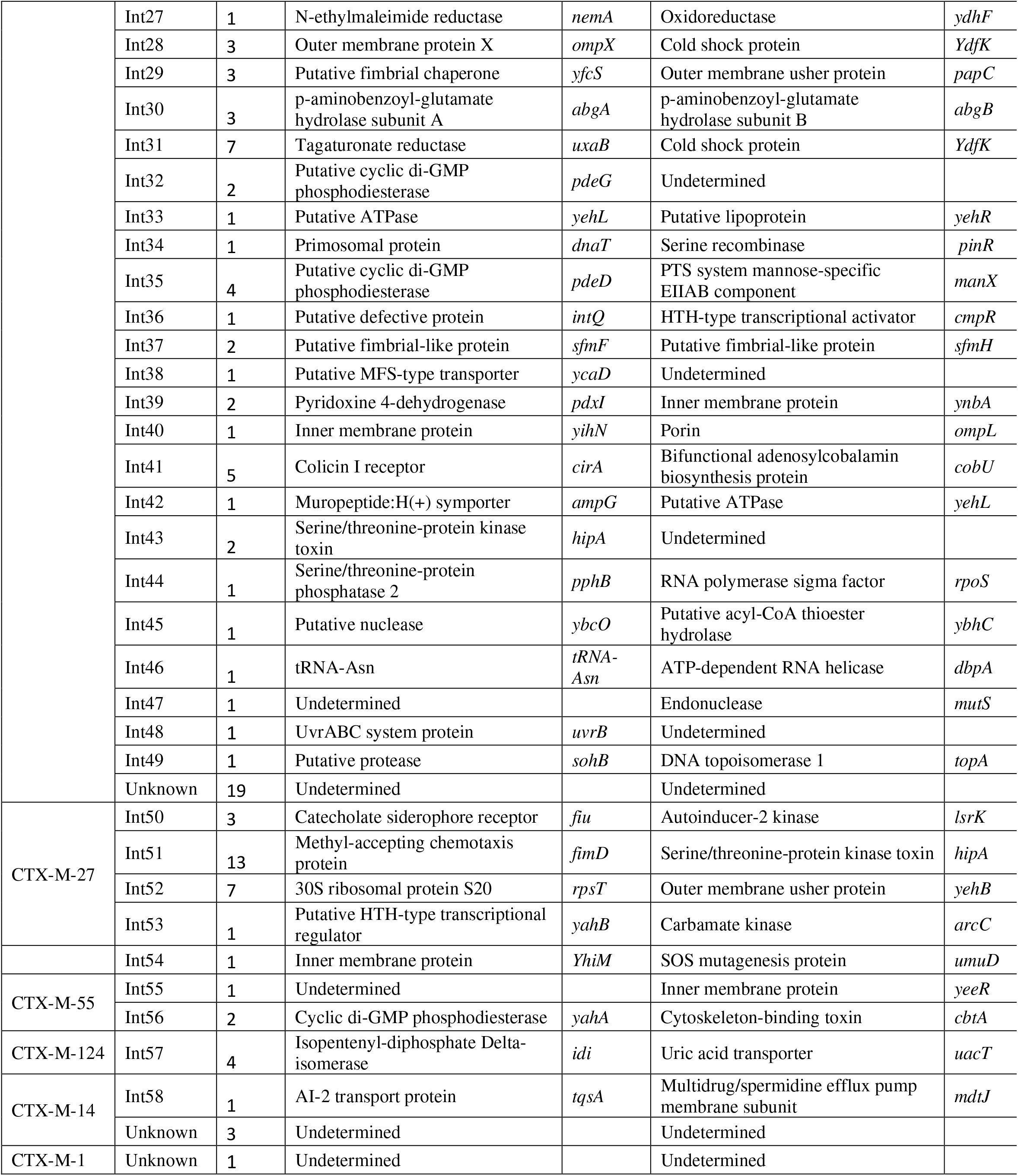
Integration sites of chromosomal *bla*_CTX-M_ genes. (A) Location of *bla*_CTX-M_ integration sites on the *E. coli* K12 linkage map. The total length of the *E. coli* K12 genome is conventionally 100 minutes. The coordinates of each insertion site are expressed in minutes on the circular map. Integration sites in olive green correspond to CDS upstream and downstream of *bla*_CTX-M_ distant of less than 10kb in the *E. coli* K-12 genome. When distant by more than 10kb, integration sites (in brown) were positioned on the *E. coli* K-12 genome with respect to the upstream CDS. *oriC* – origin of replication, *terC* – termination of replication. (B) Correspondence between integration site code and CDS upstream and downstream of the *bla*_CTX-M_ gene. Undetermined – CDS not determined because of contig break.

**Figure S10.**
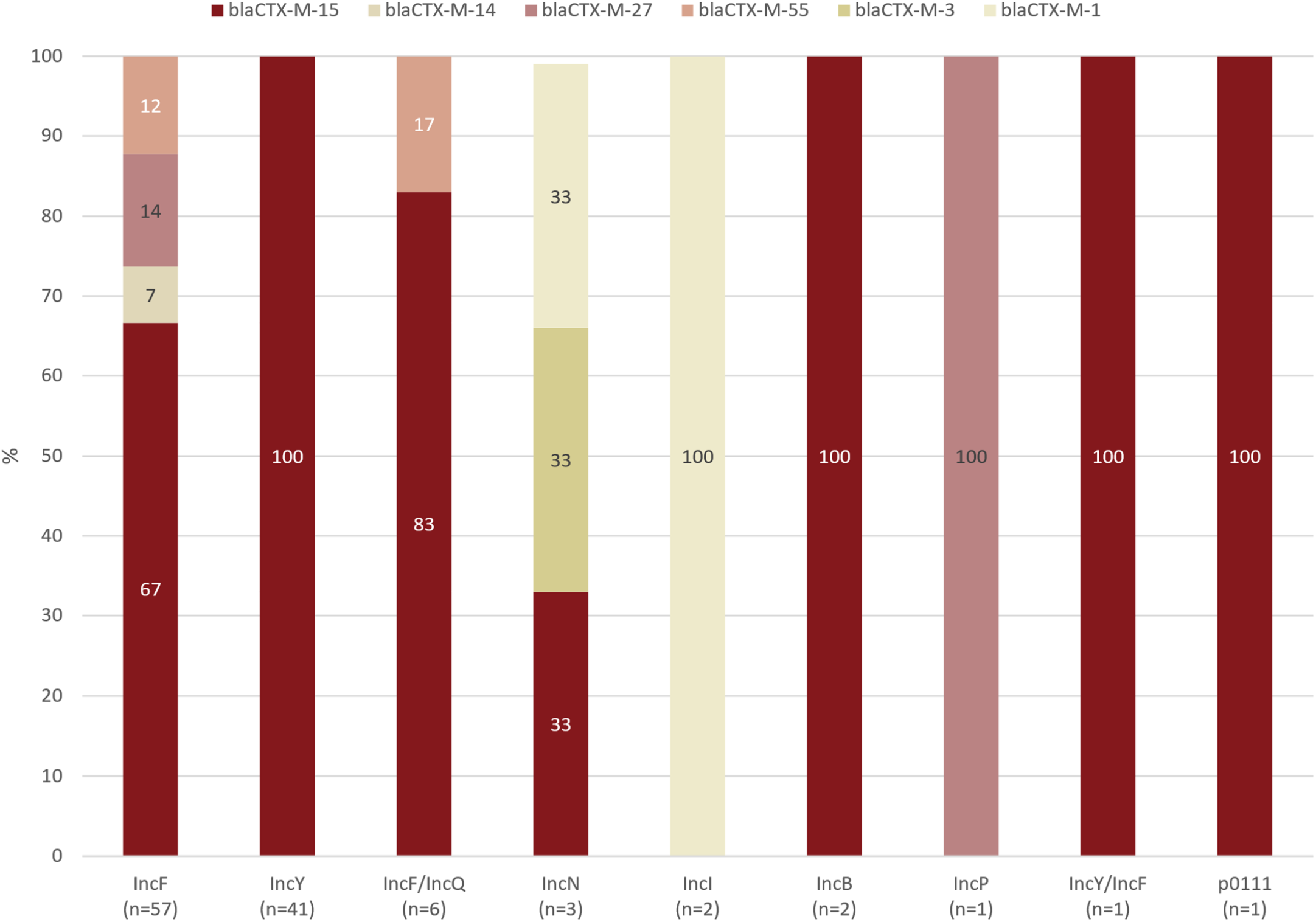
Distribution of *bla*_CTX-M_ genes among ESBL carrying plasmids.

**Figure S11.**
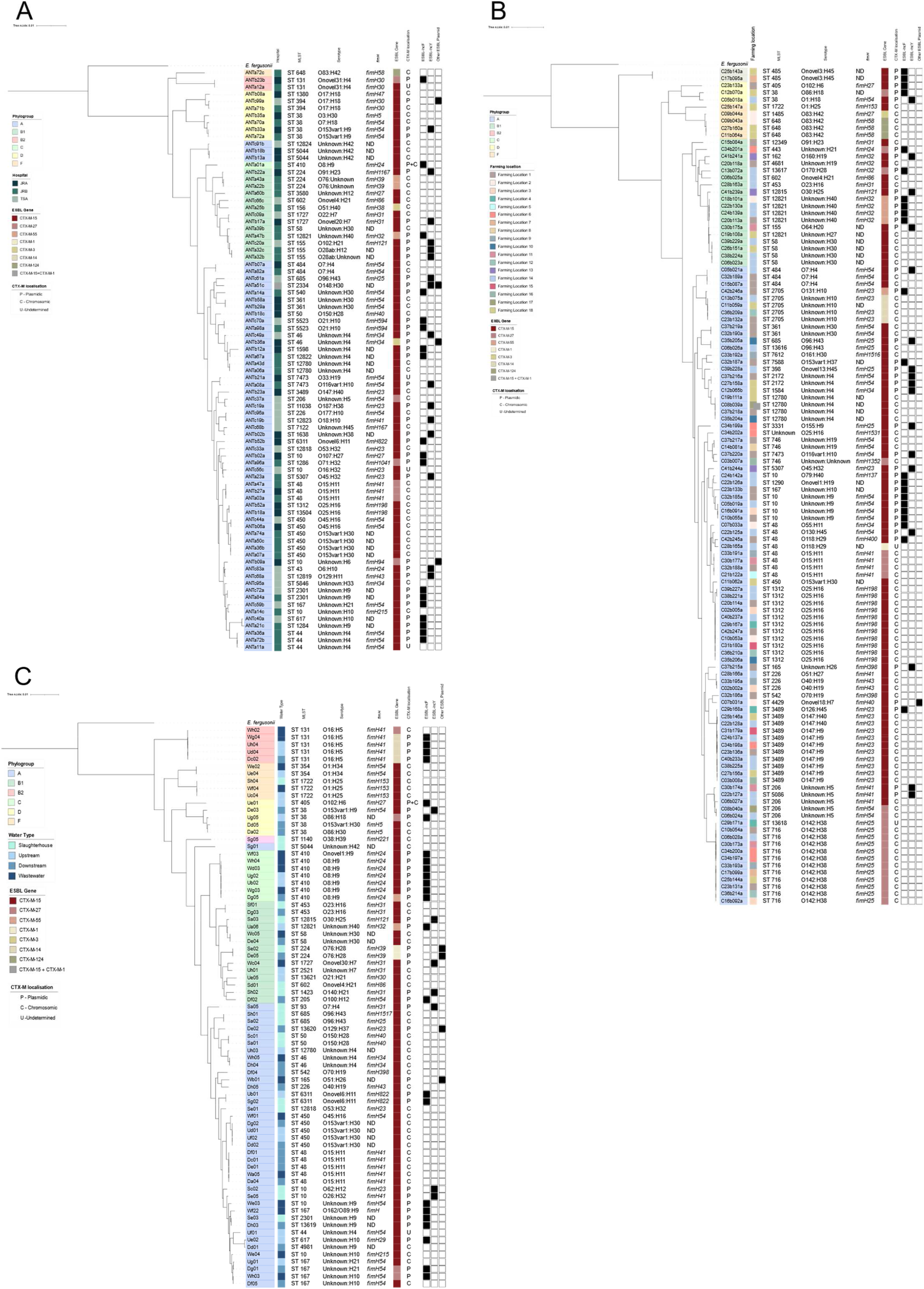
Phylogenetic trees by sector based on core genome sequences of isolates from each sector. Trees have been constructed with *E. fergusonii* used as the root. Isolate names are highlighted depending on their phylogenetic group (inner strip). Strips three to five represent isolates ST, serotype and *fimH* allele. Strips six and seven represent the *bla*_CTX-M_ gene and its location respectively. The outer strip represents the ESBL carrying plasmid replicon. **(A)** Phylogenetic tree of human isolates. The second strip represents the different hospitals (Joseph Ravoahangy Andrianavalona Hospital (JRA), Joseph Raseta Befelatanana Hospital (JRB), Mère-Enfant Tsaralalana Hospital (TSA)). **(B)** Phylogenetic tree of chicken isolates. The second strip represents the farming location. **(C)** Phylogenetic tree of environmental isolates. The second strip represents the sampling location.

**Figure S12.**
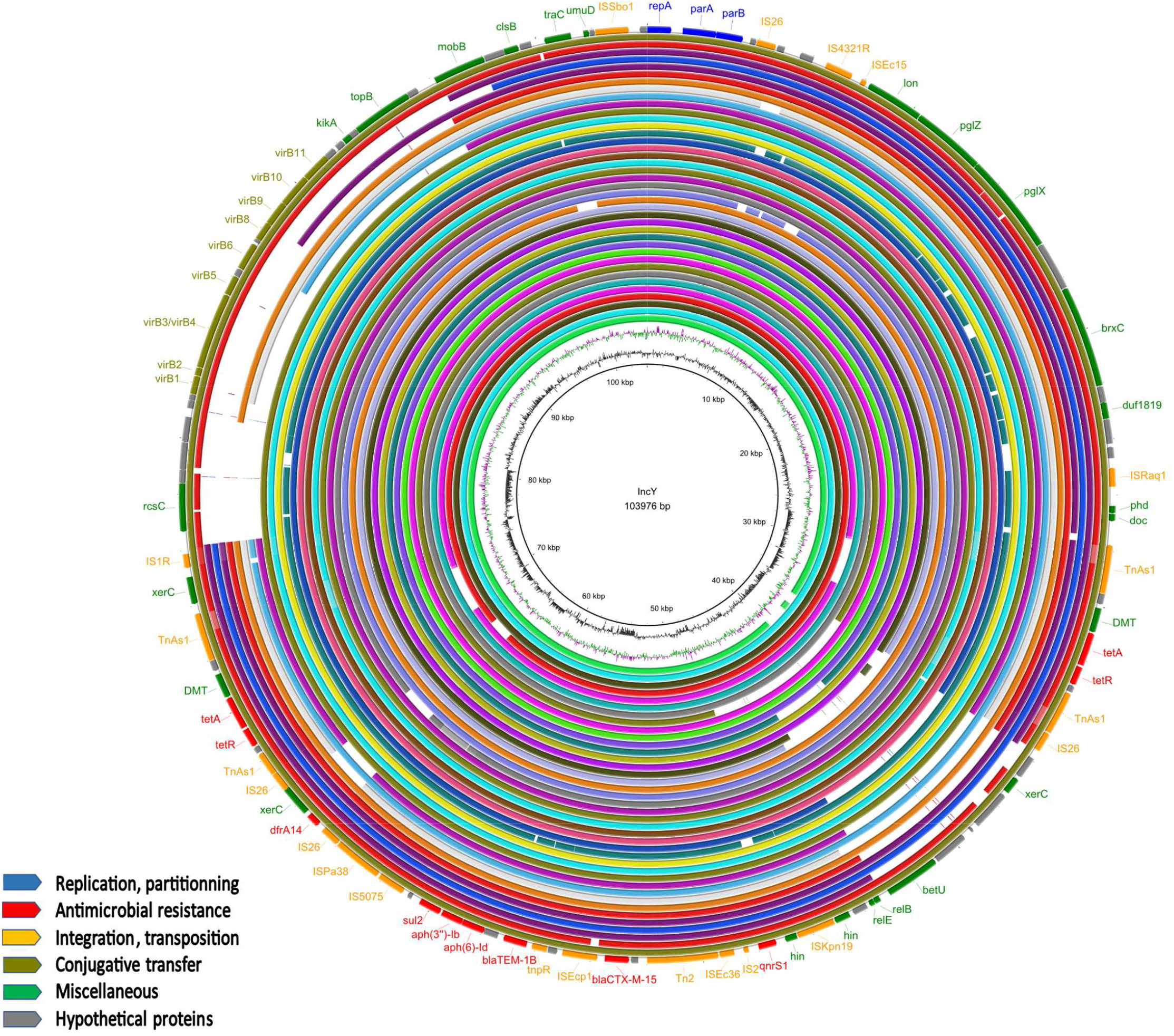
Alignment of the IncY plasmid sequences. GC content and GC skew are depicted in the inner map with distance scale. Predicted coding sequences of the reference plasmid are depicted in the outer ring with antimicrobial resistance genes in red. Two isolates were excluded from the alignment due to insufficient quality of the sequencing.

